# The Substrate of Sudden Death in Long-QT Syndrome is localized in the Epicardium

**DOI:** 10.1101/2021.11.22.21266568

**Authors:** Carlo Pappone, Giuseppe Ciconte, Luigi Anastasia, Valeria Borrelli, Edward Grant, Gabriele Vicedomini, Vincenzo Santinelli

## Abstract

Despite significant advances in the prevention of cardiovascular diseases, sudden cardiac death (SCD) persists as a major public health problem. Among young and apparently healthy individuals, Long-QT syndrome (LQTS) represents a leading progenitor of SCD owing to fatal ventricular arrhythmia. Scientific understanding of this association has grown in recent years, and the mortality rate after LQTS diagnosis has significantly decreased. However, despite medical treatment advances, life-threatening ventricular arrhythmias still occur. Until now, no research has established the degree to which this inherited condition arises from an underlying arrhythmogenic electroanatomical substrate. Here, we present direct evidence showing that LQTS patients who survive spontaneous malignant arrhythmias harbor structural electrophysiological abnormalities localized in the epicardium of the right ventricle. We further show that the elimination of these abnormalities by means of catheter ablation successfully suppresses malignant arrhythmias, offering a new approach for the effective treatment of LQTS patients.

## INTRODUCTION

Unheralded sudden cardiac death (SCD) persists as a major public health problem, claiming millions of lives every year. Most instances of SCD occur in older patients as a consequence of ventricular arrhythmias associated with ischemic heart disease. But, a great many young, apparently healthy individuals, having presented no evidence of structural heart disease, suffer instances of ventricular fibrillation (VF) ending in SCD.^1,2^ Post-mortem genetic analysis provides molecular evidence that associates as many as one-fifth of these unexplained deaths with a pathogenic basis of long-QT syndrome (LQTS),^2,3^ leading to torsades de pointes (TdP) and ventricular fibrillation (VF).^2^ Beginning with its initial description as congenital pediatric condition, LQTS has been associated with a heterogeneous family of disorders.^4^ The disease relates to mutations in more than fifteen genes,^2^ most of which encode for subunits of ion channels that lead to a prolongation of the QT interval.

One large-scale study of neonates estimates the prevalence of this disorder at about 1 in 2000 individuals.^5^ Scientific understanding has increased.^6^ But, despite advances in treatment, LQTS remains a lethal disease. Untreated genotyped but asymptomatic patients face a 36% chance of a nonfatal cardiac event, and a 13% risk of SCD.^7^ Nearly half of cardiac arrest survivors can expect VF recurrence,^2^ underscoring the fact that current therapeutic strategies fail a substantial fraction of patients known to be at risk.

In clinical practice, cases of recurrent VF requiring ICD therapy present a difficult challenge. Correctly identifying the cause of an episode constitutes a matter of life and death. Recent developments in cardiac imaging and mapping techniques have shown that the epicardial region of the heart plays a key role in the development of ventricular arrhythmias in various cardiac inherited diseases such as Brugada syndrome, arrhythmogenic cardiomyopathy and dilated cardiomyopathy.^8^ Since its first description, question has remained whether this affliction arises from an electrical manifestation or as the expression of an anatomical abnormality. With this in mind, we focused the present study on twelve consecutive LQTS patients who suffered from recurrent VF. Surprisingly, our results reveal a subtle electroanatomical substrate that served to form the mechanistic source of VF in all of these patients. This phenotypic characterization and the success of our therapeutic intervention have important clinical implications for a pathophysiological therapeutic approach in high-risk LQTS patients.

## RESULTS

### Clinical Characteristics of the Study Patients

Twelve patients (7 male, 58.3%; mean age 41.7±9.4 years, range 22-53) underwent a combined endo-epicardial mapping procedure. All patients experienced a median of four appropriate ICD discharges (range 2-8) before the procedure due to spontaneous VF episodes (Table 1). They experienced the index cardiac event at an average age of 25.3±7.9 years, whereas the mean age at the time of first ICD therapy was 32.5±6.5 years. Nine patients (75%) were taking beta-blockers, whereas in the remaining three (25%) the dose was reduced because of poor tolerability. Genetic testing revealed a *KCNQ1* mutation in three patients (25%), a *KCNH2* in three (25%), a *SCN5A* in one (8.3%) and a *KCNE1* mutation in another patient (8.3%). The genetic testing was negative in the remaining four patients (33.4%). The average QTc interval was 508.1±33.3 msec (median 518.5). None of the patients had detectable structural heart disease by advanced imaging techniques including high-resolution CT scan (4 patients) and magnetic resonance (8 patients). Programmed electrical stimulation detected inducible VF in 8 patients (66.7%). Table 1 lists the baseline clinical characteristics of the study population.

**Table 1.** Clinical characteristics of the study population.

| **N.** | **Sex** | **Initial Symptom** | **Spontaneous VT/VF Episodes at ICD recording, n** | **Time from ICD to First Episodes, y** | **Genetic mutation** | **Inducibility at EPS before ablation** | **Mode of induction** |
| --- | --- | --- | --- | --- | --- | --- | --- |
| **1** | M | CA | 5 | 32 | *KCNH2* | Y | 1 Extrastim |
| **2** | M | CA | 4 | 25 | *KCNQ1* | Y | 2 Extrastim |
| **3** | M | SYNCOPE | 2 | 10 | *KCNH2* | N | Non inducible |
| **4** | M | SYNCOPE | 6 | 18 | *KCNE1* | N | Non inducible |
| **5** | F | SYNCOPE | 2 | 29 | Negative | Y | 3 Extrastim |
| **6** | F | CA | 4 | 31 | *KCNH2* | Y | 3 Extrastim |
| **7** | F | CA | 2 | 27 | *SCN5A* | Y | 3 Extrastim |
| **8** | M | CA | 7 | 36 | *KCNQ1* | Y | 1 Extrastim |
| **9** | F | SYNCOPE | 2 | 32 | *KCNQ1* | N | Non inducible |
| **10** | M | SYNCOPE | 3 | 15 | Negative | Y | 3 Extrastim |
| **11** | F | CA | 4 | 33 | Negative | Y | 2 Extrastim |
| **12** | M | SYNCOPE | 8 | 28 | Negative | N | Non inducible |

List of abbreviations: CA cardiac arrest, EPS electrophysiological study, SD sudden death, VT/VF ventricular tachycardia/fibrillation

### Epicardial Electrophysiological Abnormalities

All twelve patients underwent combined endo and epicardial mapping. A mean of 857.4±387.8 (endocardial right ventricle [RV]), 938.2±403.7 (endocardial left ventricle [LV]) and 2684.2±1070.9 (epicardium) sites were recorded per patient. All subjects showed normal electrograms on the RV endocardium and the LV endo-epicardium (Supplemental Figure 1). Figures 1-2 and Supplemental Figures 1-9 show examples of the localization and distribution of the abnormal fragmented and/or low-voltage (<1.5 mV) signals exhibiting multiple components. These abnormal waveforms were exclusively localized over the RV epicardium, with a longitudinal distribution extending geometrically from the outflow-tract (RVOT) to the anterior wall and the infero-lateral peri-tricuspid region. Table 2 shows the electrophysiological characteristics of the study patients. Overall, the region of abnormal potential showed a lower voltage amplitude (0.89±0.23 mV) compared with the other epicardial or endocardial mapped regions (healthy RV epicardium 2.45±0.47 mV; LV epicardium 3.77±1.28 mV; RV endocardium 2.48±0.69 mV; LV endocardium 3.13±0.57 mV; p < 0.001). The epicardial signal duration of the abnormal area was also prolonged compared to the other regions (RV substrate 92.58±26.11, RV endocardium 60.99±9.41, LV endocardium 66.52±8.71, RV healthy epicardium 65.57±10.57, LV epicardium 67.46±8.85 msec; p < 0.001). The mean area with epicardial abnormalities was 14.9±3.5 cm^2^ (median 16.1 cm^2^). In the study cohort, two patients had a baseline QTc interval of 445 and 457 msec at the time of epicardial mapping, which lengthened to 524 and 584 msec after adrenaline infusion, respectively. Electrophysiological mapping was repeated under these conditions. We observed a substrate expansion of 7.8 to 16.6 cm^2^ (Supplemental Figures 4 a-b) in one patient, and of 12.8 to 17.3 cm^2^ in the second patient (Supplemental Figures 5 a-c). One of the 12 patients (8.3%) had an early repolarization (ER) pattern in the inferior leads, whereas another (8.3%) had a positive ajmaline test showing a type 1 Brugada syndrome (BrS) pattern (Supplemental Figures 6-7). Substrate was successfully eliminated in these patients with BrS and ER overlap, respectively (Supplemental Figures 6-7).

**Table 2.** Mapping and Ablation data.

| **N.** | **No. of**<br>**Abnormal**<br>**Areas** | **Location of**<br>**Abnormal Areas** | **QTc interval**<br>**before**<br>**ablation** | **Abnormal**<br>**Surface Area,**<br>**cm<sup>2</sup>** | **RF Delivery**<br>**on Substrate,**<br>**min** | **QTc interval**<br>**after ablation** | **Follow-Up** | **Inducibility**<br>**after RF** |
| --- | --- | --- | --- | --- | --- | --- | --- | --- |
| **1** | 2 | RV ant - RVOT | 505 | 16.2 | 18 | 431 | 12 | nsVF |
| **2** | 1 | RVOT | 509 | 10.7 | 11 | 483 | 26 | N |
| **3** | 1 | RV ant | 549 | 10.5 | 14 | 521 | 44 | N |
| **4** | 3 | RV ant – RV<br>inf/lat - RVOT | 535 | 20.4 | 16 | 455 | 19 | N |
| **5** | 2 | RV ant – RVOT | 539 | 11.6 | 11 | 496 | 30 | N |
| **6** | 3 | RV ant – RV<br>inf/lat - RVOT | 525 | 17.8 | 17 | 482 | 36 | N |
| **7** | 2 | RV ant – RVOT | 523 | 19 | 9 | 477 | 6 | N |
| **8** | 1 | RV ant | 445 | 16.6 | 12 | 419 | 5 | N |
| **9** | 2 | RV ant –<br>RV inf/lat | 471 | 11.6 | 9 | 369 | 31 | N |
| **10** | 2 | RV ant - RVOT | 525 | 12.1 | 12 | 372 | 34 | N |
| **11** | 3 | RV ant – RV<br>inf/lat - RVOT | 457 | 17.3 | 26 | 461 | 3 | N |
| **12** | 2 | RV ant - RVOT | 514 | 15.9 | 10 | 484 | 28 | N |

List of abbreviations. Ant: anterior; inf/lat: infero-lateral; RF: radiofrequency; RV: right ventricle,; RVOT: right ventricle outflow tract

**Figure 1.**
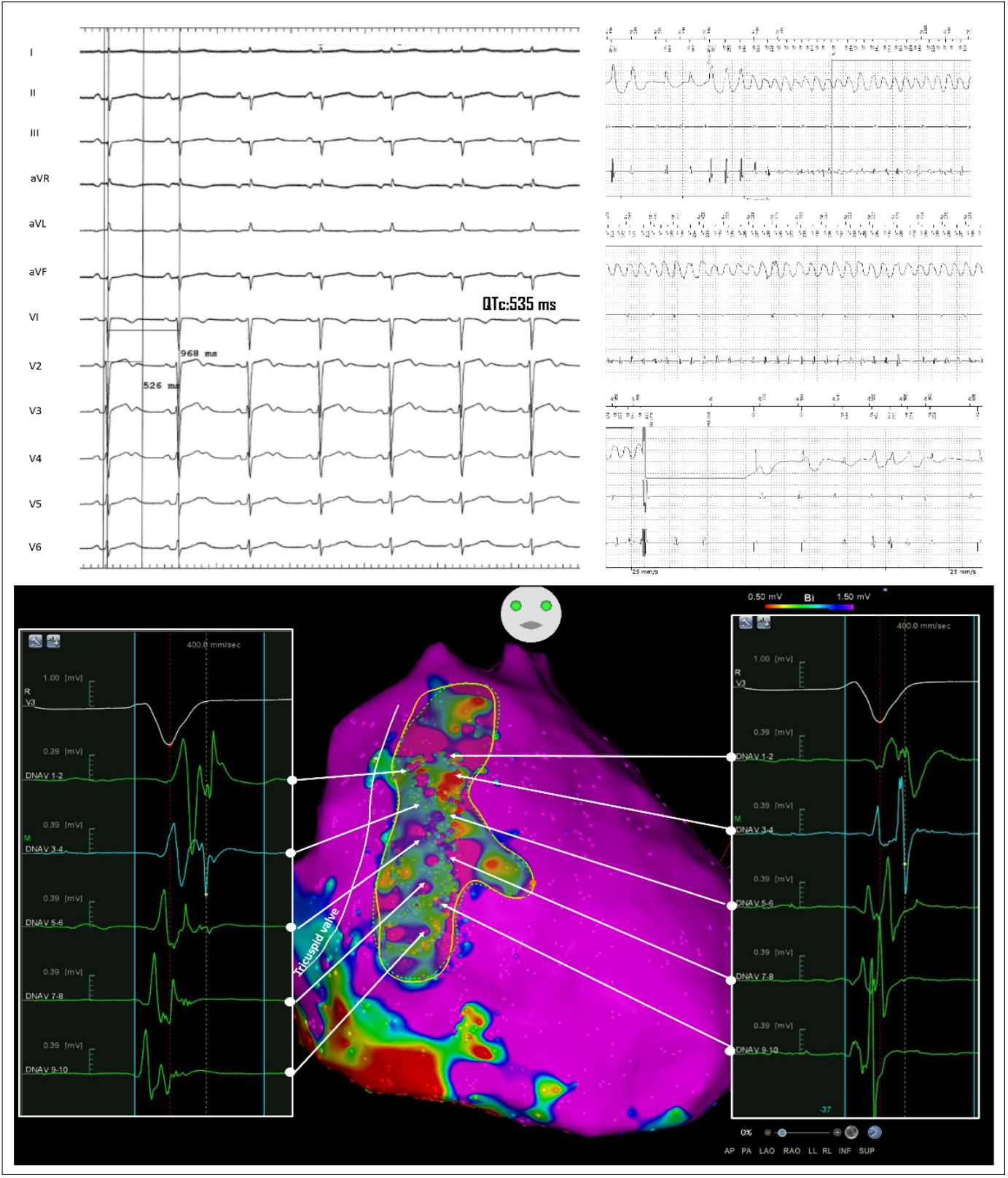
Epicardial Arrhythmogenic Substrate. Patient n°4. LQTS male patient (QTc 535 msec, ECG top panel) surviving a previous cardiac arrest and experiencing several appropriate ICD shocks (ICD recording, top panel). Epicardial mapping identified abnormal electrograms characterizing the arrhythmogenic substrate (CARTO map, bottom panel). The abnormal signals present a low amplitude (<1.5mV) and are characterized by multiple fragmented and double components (bottom panel on the left side) opposed to the normal epicardial signal which is sharp and not fractionated (bottom panel on the right, purple boxes). The distribution of abnormal signals extended from the epicardial outflow-tract to the infero-lateral peritricuspid region including part of the anterior wall. These abnormalities accounted for a surface area of 20.4 cm^2^.

**Figure 2.**
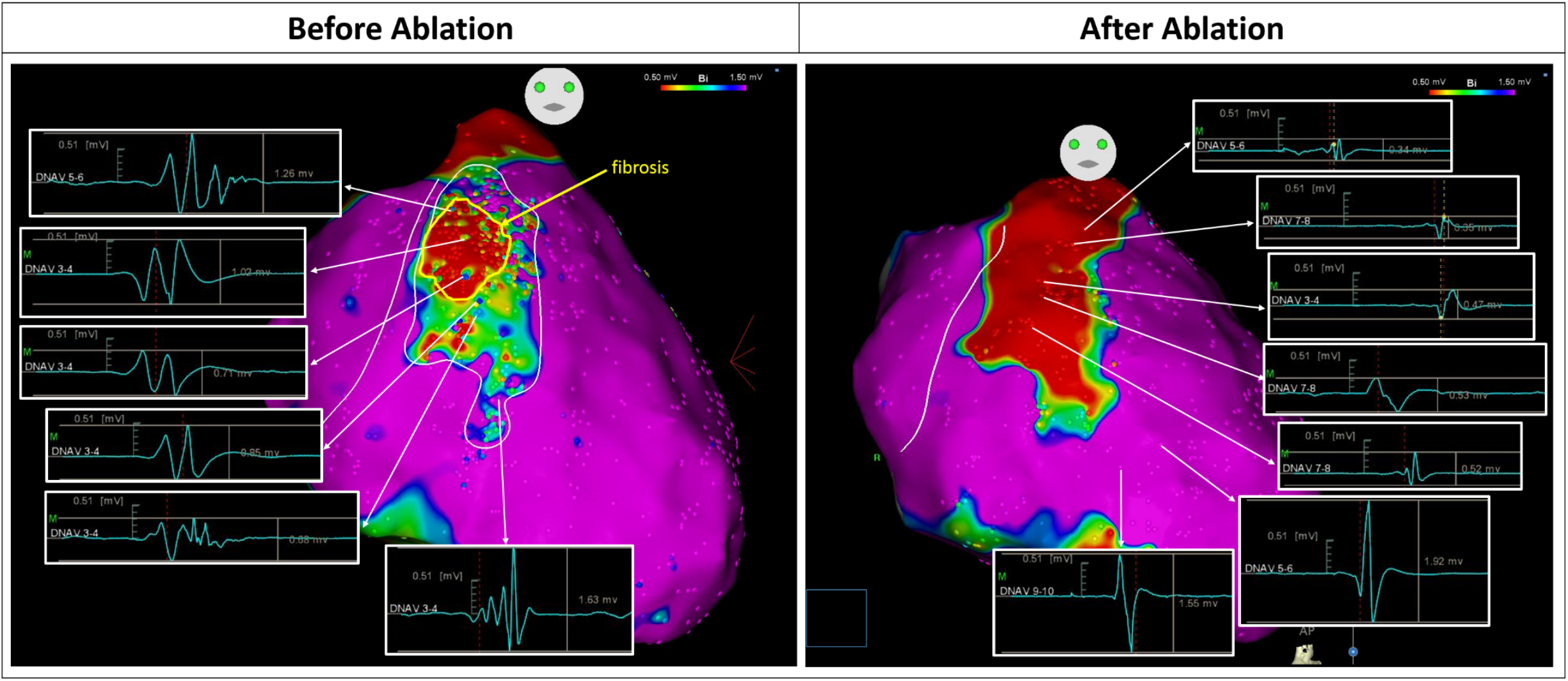
Effect of arrhythmogenic substrate elimination. Patient n°6. LQTS female patient experiencing ICD shocks. The epicardial mapping (left panel) showed a large arrhythmogenic substrate (17.8 cm^2^) involving a wide surface of the RVOT and anterior RV epicardium. The region depicted in red represents an area of low voltage potentials (<0.5 mV), indicating the progression of the disease towards fibrosis. The abnormal potentials presented fragmentation and multiple components over this area (same characteristics described in Figure 1). Catheter ablation over the arrhythmogenic substrate area resulted in the disappearance of the abnormal electrogram fragmentation (EGM example, right panel).

### Electrophysiological and Clinical Results after Epicardial Substrate Ablation

During hospitalization, only one patient showed premature ventricular contractions (PVC) and he also underwent endocardial ablation of the ectopic focus (Supplemental Figure 2a). Catheter ablation was performed over the abnormal substrate during sinus rhythm in all patients. The goal was to completely abolish electrogram fractionation in the region of interest (Figure 2 and Supplemental Figures 2b, 2d, 3b, 4b, 5b, 6 and 7). The abnormal signals were abolished after 16.4±4.1 minutes (median, 15.5 minutes) of radiofrequency application. Programmed ventricular stimulation was also performed to assess the status of inducibility after ablation. Of the eight initially inducible patients, seven became completely noninducible (87.5%) despite three extrastimuli (p=0.016); in the remaining patient (12.5%), non-sustained VF was induced only after three extrastimuli after ablation, whereas VF was initially inducible after only one extrastimulus. Upon follow-up, no spontaneous VF occurred after ablation in this patient.

No severe periprocedural complications were documented. One patient developed pericardial effusion two weeks after the procedure, possibly related to the extensive ablation (> 15 cm^2^ substrate area ablated). He underwent successful pericardiocentesis and received colchicine and nonsteroidal anti-inflammatory drugs without recurrence of pericarditis. At discharge, 12-lead ECG confirmed a significant shortening of the QTc interval (≥ 40 msec) in 7/12 (58.4%) patients (Figure 3), with a mean QTc duration of 454.2±47.7 msec (median 469 msec) compared with the pre-procedural 508.1±33.3 msec (p < 0.001). After a median follow-up of 22.8±13.6 months, no VA recurrences were detected during ICD monitoring (p=0.002).

**Figure 3.**
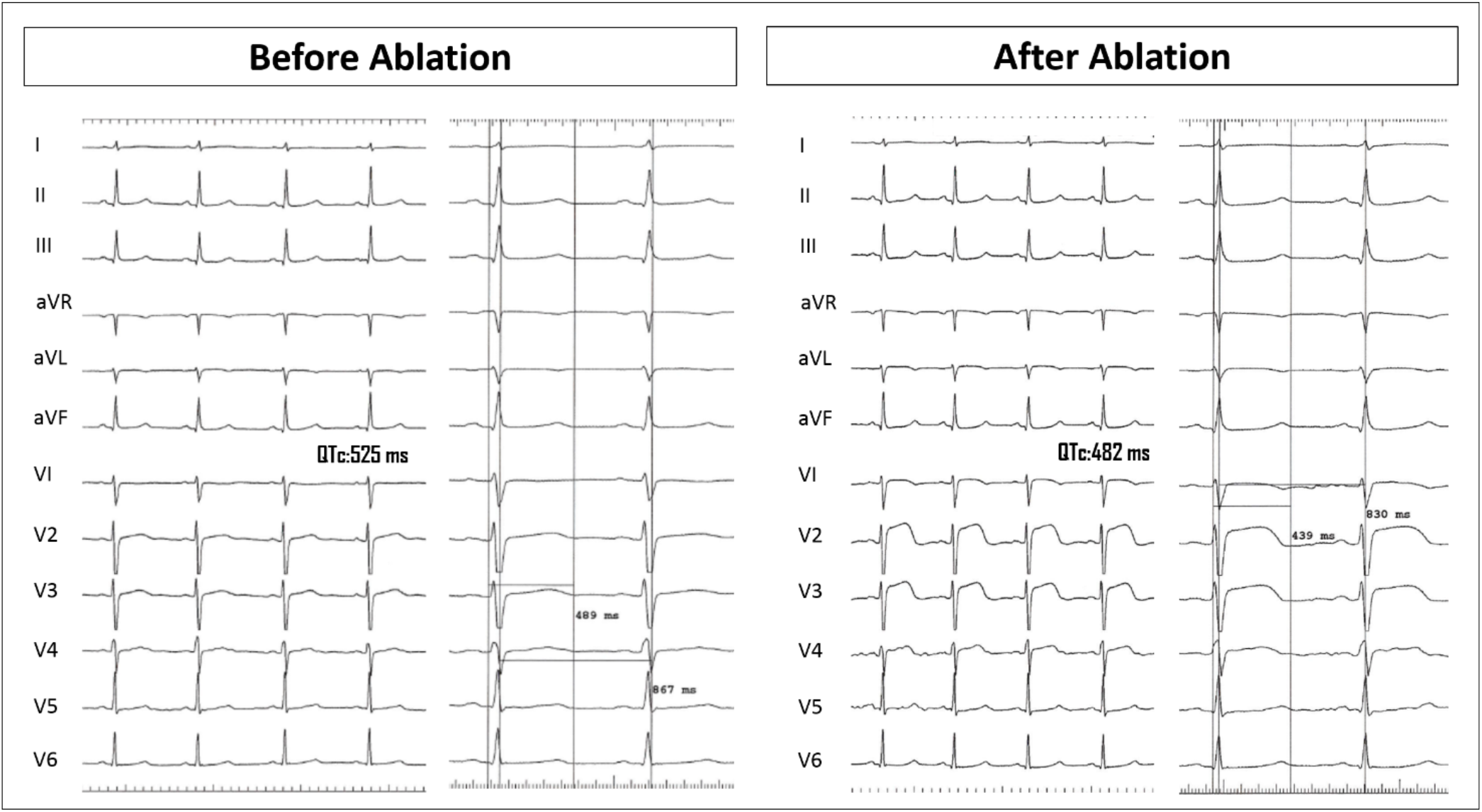
ECG changes before and after ablation. The 12-lead ECGs (same patient as in Figure 2), recorded at the beginning (left panel) and end (right panel) of the procedure, show QTc shortening (from 525 to 482 msec). In the right panel, transient ST-segment elevation is observed in V1-V3 lead due to the epicardial ablation over the anterior RV epicardium, which completely solved during the follow-up (see Supplemental Figure 8). The ECGs are shown at 25 and 50 mm/sec paper speed.

## DISCUSSION

This study provides new insight on the presence of electrophysiological abnormalities in LQTS patients who survive spontaneous malignant arrhythmias. The study cohort showed clear evidence of local electroanatomical abnormalities that were present in all treated subjects. This abnormal electrophysiological substrate is characterized by regions of low-voltage, delayed signals with multiple fragmented components localized exclusively in the epicardial layer of the right ventricle. Catheter ablation of these regions prevented recurrence of ventricular fibrillation in all cases. These findings provide evidence that localized epicardial structural abnormalities may underlie a significant subset of high-risk LQTS patients, and this region may serve as a target for ablation treatment.

Although LQTS mortality is decreased significantly in risk-stratified and treated patients, cardiac events (syncope and ICD shocks) continue to occur in nearly 25% of previously symptomatic patients despite maximal therapy,^9^ underscoring the need to improve the management of high-risk patients. It is generally believed that LQTS is a purely electrical disorder. Although intriguing proof of inhomogeneous ventricular mechanics has been reported,^10,11^ the literature lacks evidence indicating the presence of electrophysiological abnormalities in LQTS patients.

Our study establishes the presence of abnormal potentials localized exclusively in the epicardium of the right ventricle in a subset of high-risk LQTS patients. This arrhythmic syndrome relates to a diverse group of inherited disorders arising from mutations of ion channels.^12^ These fundamental alterations increase the likelihood of early afterdepolarizations triggering TdP and VF.^13^ Elimination of focal triggers by endocardial ablation has been described as a possible treatment option.^14^ Unfortunately, only one patient in our series underwent successful PVC ablation at the endocardial site. This might well reflect the fact that frequent PVCs are rarely mapped in patients with LQTS, owing in large part to concurrent drug therapy.^14^ But one must eliminate this trigger before targeting the arrhythmogenic substrate.

Once triggered, a myocardial substrate with spatially inhomogeneous conduction properties acts to maintain malignant arrhythmias.^15,16^ Prolongation of the action potential leads to a local conduction blockade at the epicardial-myocardial interface, resulting in reentrant excitation.^17^ This may explain the arrhythmogenic role of the abnormalities described above. The poorly understood nature of these local electrophysiological effects invites speculation. A traditional interpretation assigns fractionated electrograms to functional disturbances or microstructural heterogeneities favoring reentry,^18-21^ with a demonstrated role in various diseases.^22,23^ By endo-epicardial mapping, we have found that these local electrophysiological abnormalities are not transmural, but involve only the thin epicardial layer of the right ventricle, which may explain why advanced imaging techniques often miss such abnormalities, as occurred in this experience.^24^ In fact, these microstructural abnormalities might lie under the spatial resolution threshold of currently available imaging techniques, especially when assessing the thin RV wall. In such cases, *electroanatomical imaging* using tridimensional mapping techniques may well serve as a complementary tool integrating the information of other approaches, to adequately define the arrhythmogenic substrate, not only in patients with structural heart diseases, but also in subjects with ostensibly normal hearts.

In this population, the autonomic nervous system (ANS) and increased sympathetic activity play an important role in the mechanism of malignant arrhythmias.^25^ The heart is innervated by an extremely complex intrinsic and extrinsic cardiac ANS that regulates the electrophysiological properties of cardiomyocytes.^13^ Although this is not the case in all LQTS types, an increased adrenergic tone may prolong the QT interval, setting the stage for heterogeneity of repolarization. Moreover, the abnormal substrate localization might correspond to the anatomical distribution of ANS on the RV, as sympathetic fibers are predominantly localized in the subepicardium (Figure 4).^26,27^ As a result, imbalanced innervation density and chronically increased adrenergic tone can lead to structural and functional remodeling,^13^ subtle dysfunction, and myocardial damage, resulting in locally abnormal potentials that lead to fatal arrhythmias, as seen in heart failure.^26^

**Figure 4.**
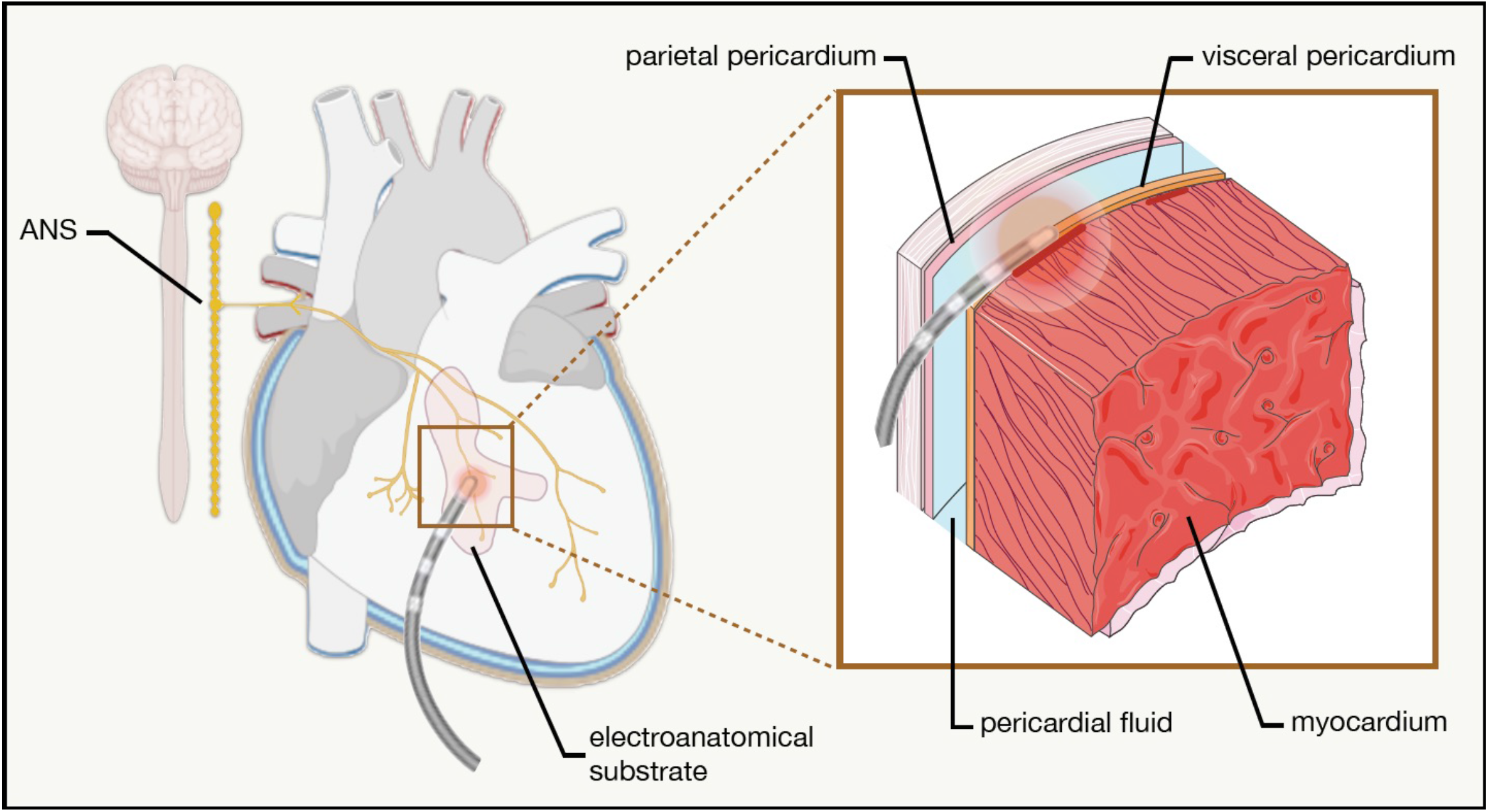
Epicardial ablation of LQTS electroanatomical substrate. Schematic representation of the distribution of the orthosympathetic nervous system (ANS) in the epicardial layer of the right ventricle. Ablation of the abnormal area (LQTS electroanatomical substrate) is performed by epicardial access. A schematic representation of the ablation procedure is shown in the magnified panel.

Recent progress in the mapping of cardiac arrhythmias has revealed the crucial role played by the epicardial region in the activation of arrhythmogenic mechanisms. In fact, documented electroanatomic abnormalities have already established the right ventricular epicardium as an area of interest in several inherited cardiomyopathies (e.g., Br and ER syndromes, arrhythmogenic right ventricular cardiomyopathy, and idiopathic VF).^28-32^

In light of these findings, the epicardium has emerged as an important determinant in SCD-related cardiomyopathies, evolving from a neglected region to a virtual “ *fifth*” chamber of the heart. Similarities of cause and effect suggest a common network of genetic architecture and phenotypic expression at the root of many, apparently distinct diseases. The broad clinical spectrum of these disorders may also overlap,^33-35^ suggesting a possible pathophysiological link in a specific subgroup of patients yet to be identified.

Thus, increasing evidence suggests that inherited diseases with grossly normal hearts should be considered cardiomyopathies because ion channel dysfunction affects cardiomyocytes function and structure.^35^ Consistent with these concepts, two patients in the present study population exhibited an overlapped manifestation of LQTS with patterns of ER and BrS, including a detection of the arrhythmogenic substrate associated with ER and BrS.^28,30,36^ These manifestations differed mainly in the localization and distribution of epicardial abnormalities (Supplemental Figure 9). While ER Syndrome tends to localize in the epicardium of both ventricles,^30^ BrS shows a confluent and homogeneous ‘onion-like’ substrate in the right ventricle.^28^

All of patients in this study exhibited consistent electroanatomical abnormalities, distributed longitudinally from the epicardial RVOT to the anterior peri-tricuspid region, further supporting the pathophysiological link between these entities as epicardial cardiomyopathies. In particular, our observations do not support the speculation that other coexisting or concomitant diseases produce this arrhythmogenic substrate. An inflammatory response may cause widespread or scattered damage, but would be unlikely to localize in every case at the identical site and with an equivalent spatial distribution.

This pathological cause and effect may explain the benefit afforded by catheter ablation, as documented in the present study. Indeed, removal of the abnormal epicardial substrate (average 15 cm^2^) resulted in suppression of malignant ventricular arrhythmias during follow-up, providing evidence that these abnormal regions may exert an arrhythmogenic effect in LQTS patients.

This disease also carries a significant risk for life-threatening events after the fourth decade of life,^37^ which explains why many patients aged > 40 years comprise this study cohort. One could also speculate that a certain subgroup of patients develops local electrical remodeling over time due to epicardial reactivation^38^ promoting disease progression to a more severe condition.

The epicardial region presents a complex histological architecture. It contains multipotent progenitors that can differentiate into smooth muscle cells, fibroblast, adipocytes and cardiomyocytes.^8^ In addition, the epicardial adipose tissue, which differs from the paracardic adipose tissue located at the outer surface of the pericardium, is intimately connected with the subepicardial myocardium without barriers, such that peptides and adipokines can freely diffuse.^8^ The adipose tissue secretes several cytokines and adipokines,^39^ regulating the cardiac electrical properties directly and indirectly.^40,41^ The epicardium has also important signaling functions and it can be reactivated becoming a source of myofibroblasts and of growth and of angiogenic factors.^42^ Accordingly, the reactivation of the epicardium and the capacity of progenitor cells to differentiate into fibroblasts or adipocytes can lead to fibrous and/or fatty infiltrations to the subepicardial myocardium.^43^ Such microstructural electrical remodeling may well give rise to an electroanatomical fragility of the subepicardial layers that affects the local conduction properties and consequently the normal endo-epi repolarization gradient.^44^ These observations support the concept that several mechanisms induce progressive electroanatomical damage towards fibrosis in a certain subset of patients leading to potentially lethal arrhythmias (Figure 2).

The prolonged QT interval normally appears in all ECG leads. Surprisingly, we report that a QT shortening occurred in the majority of patients after elimination of the electrical abnormalities. Despite some fluctuations, the stable QT shortening invites speculation that ablation over these abnormal regions causes a distal denervation that impairs repolarization of the entire heart, thus supporting its arrhythmogenic effect. These findings demonstrate that localized epicardial structural abnormalities may underlie a significant subset of high-risk LQTS patients, and this region could serve as a target for ablation treatment, particularly if no trigger could be demonstrated.

We recognize the potential limitations of this study. One could point to its small sample size. However, we decided to focus on symptomatic patients requiring effective treatment to prevent recurrent malignant arrhythmias and ICD discharges. This enabled us to use a consistent mapping procedure and ablation strategy for all patients, adding weight to our conclusions. The mechanism of arrhythmogenesis and the correlation between genotype and clinical phenotype may vary in different types of LQTS. Therefore, these results may not apply to all subgroups of LQTS. We could not rule out the possibility that left cardiac sympathetic denervation (LCSD) may have added benefit for our patients. However, it should also be acknowledged that our approach allowed us to identify a target for catheter ablation without the potential complications of surgery.

Finally, this study demonstrates successful salvage therapy in the treatment of high-risk patients with LQTS who would otherwise be doomed to death owing to recurrent malignant ventricular arrhythmias. In this population, regions localized in the epicardium of the right ventricle exhibit structural electrophysiological abnormalities. Catheter ablation of these regions resulted in successful suppression of malignant arrhythmias that threaten sudden cardiac death, providing a novel approach for the treatment of LQTS patients. A confirmation of these promising results awaits future studies involving broader and multicenter LQTS populations.

## METHODS

### Study Population

All patients with LQTS were referred for the management of frequently recurring spontaneous VF requiring ICD shocks. We prospectively enrolled 13 patients with LQTS who had a previous history of cardiac arrests and ICD shocks owing to VF, that recurred despite antiarrhythmic therapy with β-blockers, negatively affecting quality of life of these individuals. We excluded one person because of a neurological anoxic condition following index cardiac arrest. All patients fulfilled criteria for LQTS,^45^ having undergone cardiac testing including echocardiogram, high-resolution CT/ magnetic resonance imaging, and cardiac catheterization to exclude structural heart disease. All patients signed an informed consent approved by our Institutional Review Board. All authors had full access to all data in the study and take responsibility for its integrity and data analysis.

### Mapping Procedure

None of the patients had prior mapping or ablation procedures. We performed high-resolution endocardial and epicardial electroanatomical maps for all patients by means of a 3-dimensional (3D) mapping system (CARTO 3, Biosense Webster, Diamond Bar, CA, USA) using high-density decapolar mapping catheter (DecaNAV, Biosense Webster). We recorded and analyzed data simultaneously with regard to amplitude, duration, relation to surface QRS. A subxyphoidal puncture provided epicardial access, as previously described.^46^ Detailed methods are described in the supplemental materials.

During sinus rhythm, the voltage maps were assessed using conventional voltage criteria for healthy tissue to set standard cut-off values.^29,31^ Electrogram criteria matched with those defining ischemic or dilated cardiomyopathies abnormalities.^21,47-49^ We found abnormal signals associated with fragmented low-frequency ventricular potentials, distinct from the near-field ventricular electrogram.

We classified abnormal signals in this way if they met at least one of the following criteria: 1) a wide duration (>80 msec) with multiple fragmented components (>3 distinct peaks); 2) a component of low amplitude <1.5 mV; 3) a distinct and delayed component beyond the end of the QRS complex, 4) multiple deflection activities. All maps were obtained at baseline conditions and, when necessary, after drug challenge with adrenaline to achieve further QTc prolongation and to evaluate the whole extent of epicardial abnormalities. Adrenaline was infused and assessed according to Ackerman et al.^50^

### Radio-Frequency Catheter Ablation

We performed catheter ablation with the aim of eliminating all abnormal epicardial electrical activity, using an irrigated–tip ablation catheter (Thermocool SmartTouch SF, Biosense Webster) (Figure 4). Each RF application lasted 5-30 seconds, and it was delivered in a point-by-point fashion to achieve a complete abolition of the targeted potentials using a power control mode (35-50 W).

### Genetic Testing

To determine the genotype, all patients were screened for variants in LQTS-associated genes (*AKAP9, ANK2, CACNA1C, CALM1, CALM2, CALM3, CAV3, KCNE1, KCNE2, KCNH2, KCNJ2, KCNJ5, KCNQ1, SCN1B, SCN4B, SCN5A, SNTA1 and TRDN)* using genomic DNA and processed with Next Generation Sequencing (TruSight One sequencing kit with NextSeq platform). Those who were genetically negative were screened for the abovementioned variants.

### Statistical analysis

Data are presented as mean ± standard deviation or as absolute values and percentages where appropriate. The exact McNemar test was used to analyze the proportion differences of VF episodes before and after ablation (both inducible at programmed stimulation or spontaneous). The Wilcoxon test was used to compare the number of arrhythmic episodes and the QTc interval changes before and after ablation. Non-parametric analysis by the Kruskal-Wallis test was used to compare the differences of the electrogram voltage and duration among different endo-epicardial regions. P-value <0.05 was considered statistically significant. Statistical analyses were conducted using SPSS (v.21, IBM SPSS Statistics).

## Data Availability

The data that support the findings of this study are available from the corresponding author, CP, upon reasonable request.

## Acknowledgements

This work was partially supported by Ricerca Corrente funding from the Italian Ministry of Health to IRCCS Policlinico San Donato.

## Author contributions statement

C.P, G.C. and G.V. performed all the procedures and took care of patient’s management. V.B. organized and extracted digital information, electrocardiograms and had care of the database. All authors provided critical interpretation of the findings and possible pathophysiological explanations and took active part in writing and revising this manuscript.

## Data Availability Statement

The data that support the findings of this study are available from the corresponding author [C.P.] upon reasonable request.

