## Supplemental Materials for "The Substrate of Sudden Death in Long-QT Syndrome is localized in the Epicardium"

### METHODS

#### Arrhythmogenic substrate mapping

None of the patients had prior mapping or ablation procedures. All procedures were performed under general anesthesia. Blood pressure was constantly monitored through a radial arterial line. Surface ECG and intracardiac electrograms were continuously recorded and stored on a recorder system available in the EP laboratory (Claris Workmate, Abbott). Programmed ventricular stimulation was performed as previously described to assess inducibility of VF [1].

After having performed endocardial (Endo) electroanatomical mapping of the right and left ventricles (RV and LV), epicardial (Epi) access was obtained using fluoroscopy-guided subxyphoidal puncture, and a steerable sheath (Agilis EPI, Abbott, MN, USA) was introduced. Detailed Endo-Epi mapping was performed using a 3-dimensional (3D) mapping system (CARTO 3, Biosense Webster, CA, USA) with a high-density mapping catheter (DecaNAV, Biosense Webster; 1 mm electrodes with 2-8-2 interelectrode spacing). Endocardial mapping always preceded epicardial mapping, in order to obtain a correct delimitation of the endocardial boundaries when mapping the epicardium. Bipolar electrograms were filtered from 16 Hz to 500 Hz, displayed at 400 mm/s speed, and were recorded between the electrode pair. Electrogram acquisition was performed only if the multipolar catheter was stable in each epicardial position and if the EGM morphology, evaluated by the operators, was consistent and repetitive for at least 3 consecutive beats, thus avoiding artifacts. Acquisition was excluded if their technical quality was insufficient. The abnormal signals were considered form the near-field ventricular electrogram, which may display fractionation, double or multiple components separated by very-low-amplitude signals or an isoelectric interval. Total signal duration was measured for each potential and measurements were interpreted and validated online by three expert electrophysiologists using CARTO3 system electronic calipers.

**Radio-Frequency Catheter Ablation**

Areas showing abnormal EGMs were mapped with a greater point density to delineate the extent of such regions of interest. Once epicardial areas were identified and quantified, radiofrequency catheter ablation (RFA) was then extensively delivered over the abnormal regions o abolish all the abnormal electrograms. After having completed RFA of the arrhythmogenic substrate, a remap focusing on the low-voltage area was obtained after ablation to confirm the complete elimination of abnormal electrograms. The remapping was repeated following the same criteria as the initial mapping procedure. Residual abnormal electrograms were targeted with the same approach used during initial mapping and ablation. At the end of the procedure, when all the abnormal fragmented signals were successfully eliminated, programmed ventricular stimulation from the RV apex was performed to assess ventricular arrhythmia inducibility after ablation, using 3 different cycle drives (600, 500 and 400 ms) up to 3 extrastimuli with a minimal coupling interval of 200 ms or until refractoriness. End point of the procedure was the complete abolition of all fragmented abnormal potentials.

**Periprocedural care and Follow-up**

At the end of the procedure, venous and arterial sheaths were withdrawn immediately, whereas complete pericardial fluid removal and subsequent intrapericardial steroid (triamcinolone acetate 2 mg/kg) was injected[2]. Patients were continuously monitored at least 6 days in the hospital after the procedure. Patients were followed up at 3, 6 and 12 months after the procedure for clinical review and device interrogation.

### SUPPLEMENTAL FIGURES

**Supplemental Figure 1.** Example of biventricular mapping showing normal amplitude and lack of fragmentation of the bipolar endocardial electrograms.

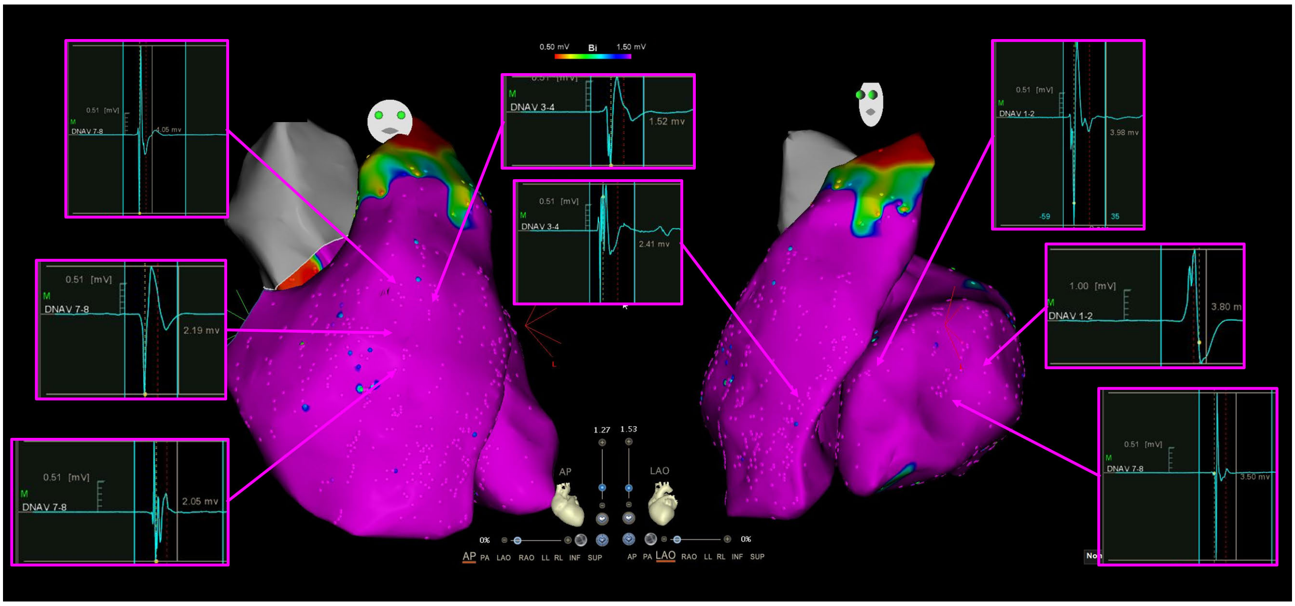

**Supplemental Figure 2a.** Same patient as in Figure 1 (patient n° 4) of the main text. LAT CARTO map showed the earliest activation site of the PVC arising from the anterior aspect of the RVOT. Signal acquisition during the spontaneous PVC is shown in the right panel.

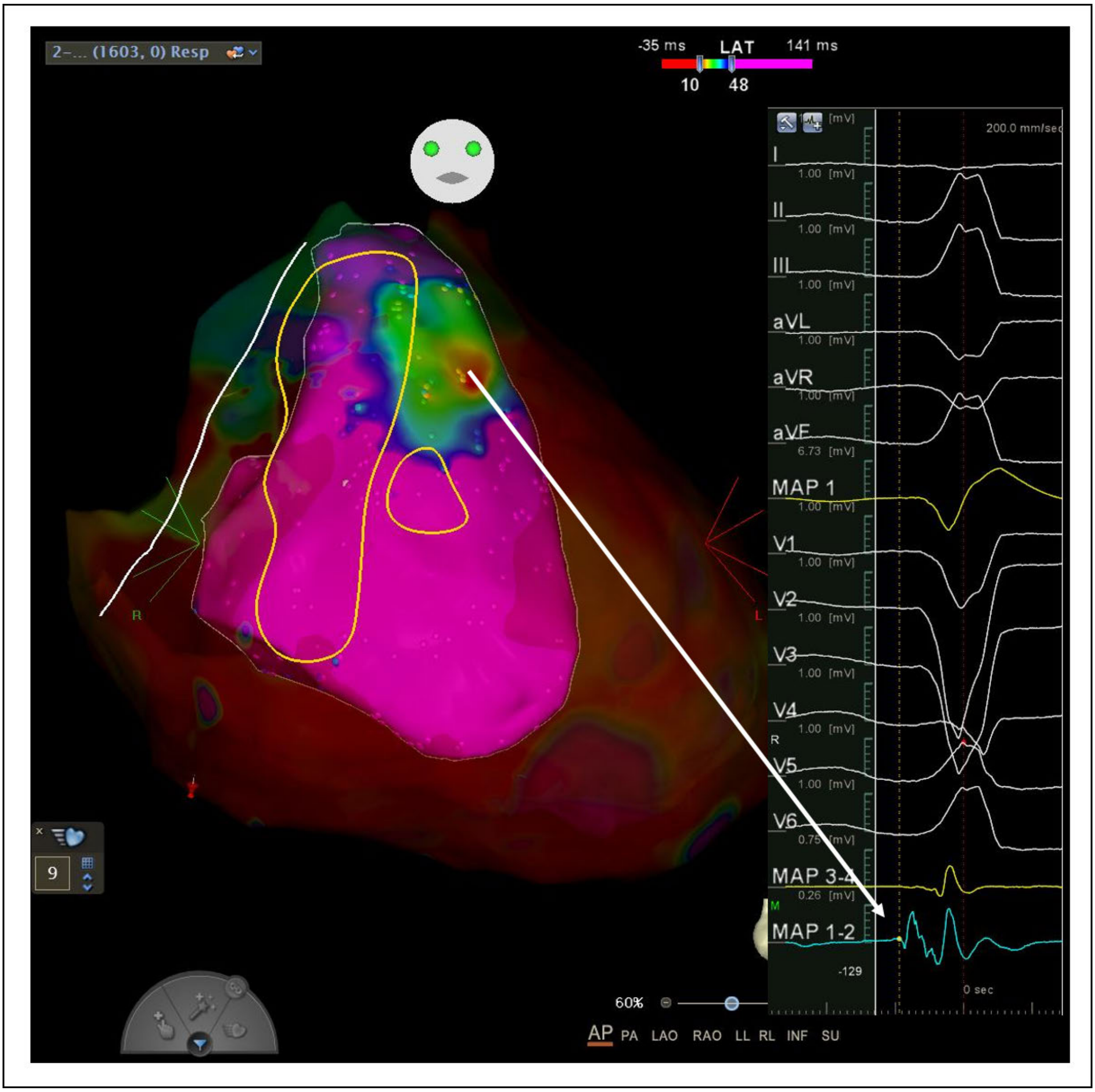

**Supplemental Figure 2b.** Same patient as in Figure 1. The 12-lead ECG (left panel), recorded at the and end of the procedure, shows QTc shortening (455 msec). The ECGs is shown at 25 mm/sec paper speed. Catheter ablation over the arrhythmogenic substrate area resulted in the disappearance of the abnormal fragmentation (EGM example, right panel). Transient ST-segment elevation is acutely observed after ablation.

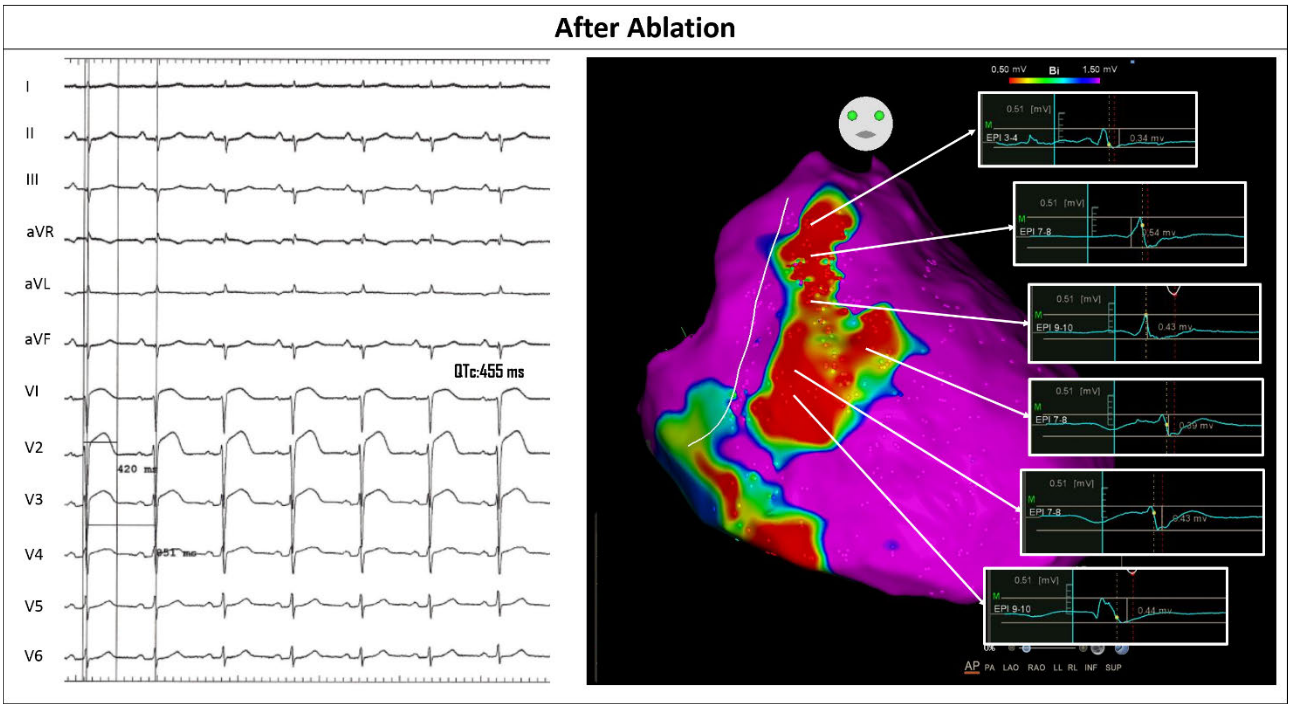

**Supplemental Figure 2c.** Same patient as in Figure 1. The 12-lead ECGs, recorded at the beginning (left panel) and end (right panel) of the procedure, show QTc shortening. The ECGs are shown at 25 and 50 mm/sec paper speed to assess the interval measure.

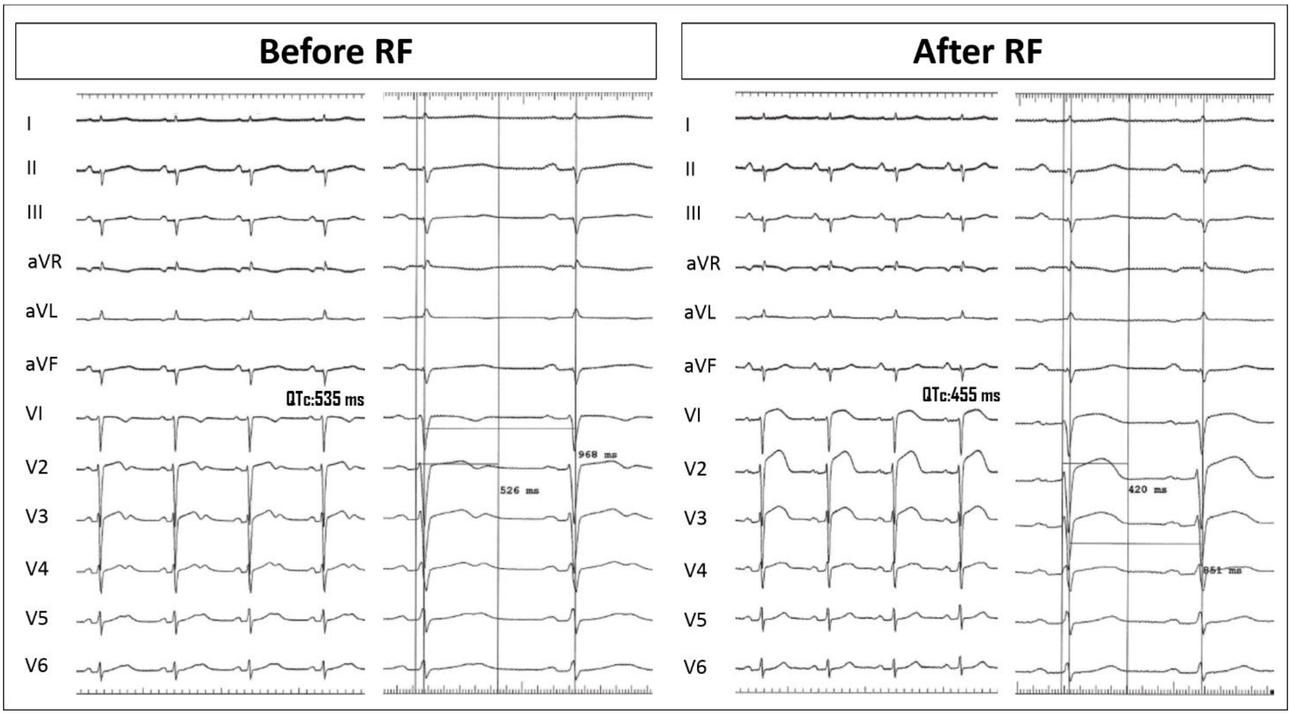

**Supplemental Figure 2d.** Epicardial mapping before (left) and after ablation (right) to assess disappearance (right panel, asterisks) of the abnormal fractionation of the electrograms with multiple and delayed components (white arrows, left panel).

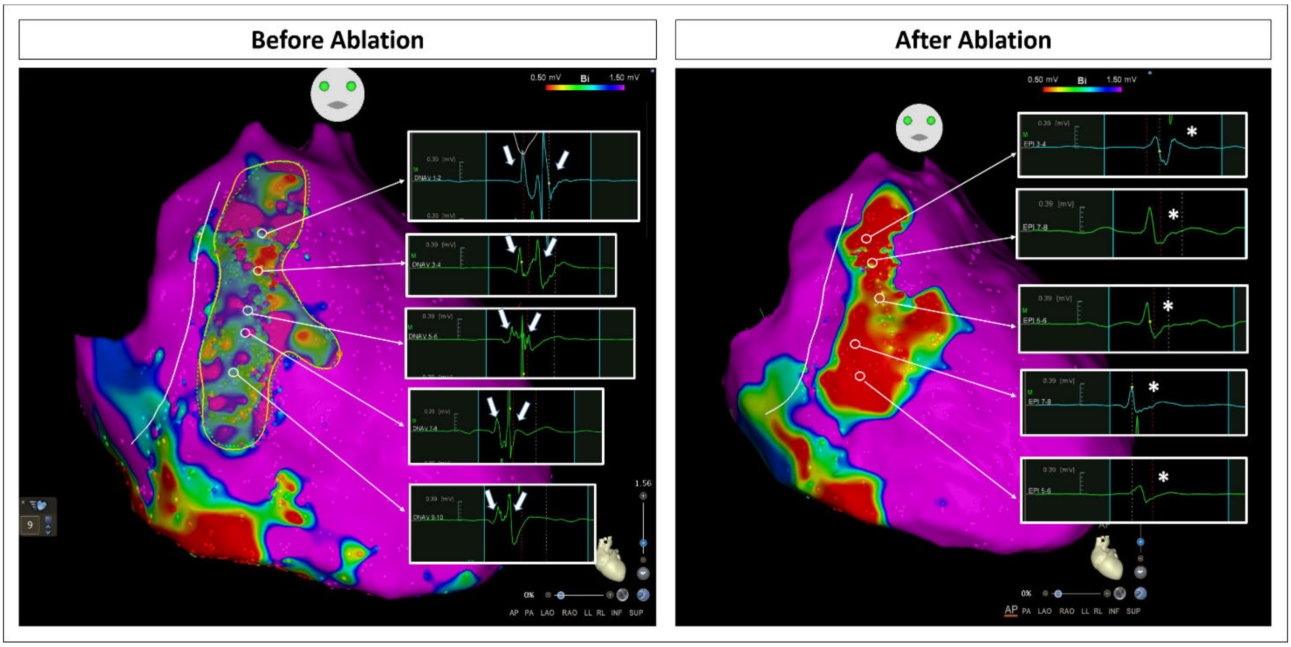

**Supplemental Figure 2e**. Long-term follow-up 12-lead ECG showing a normal QTc interval duration.

Follow-up

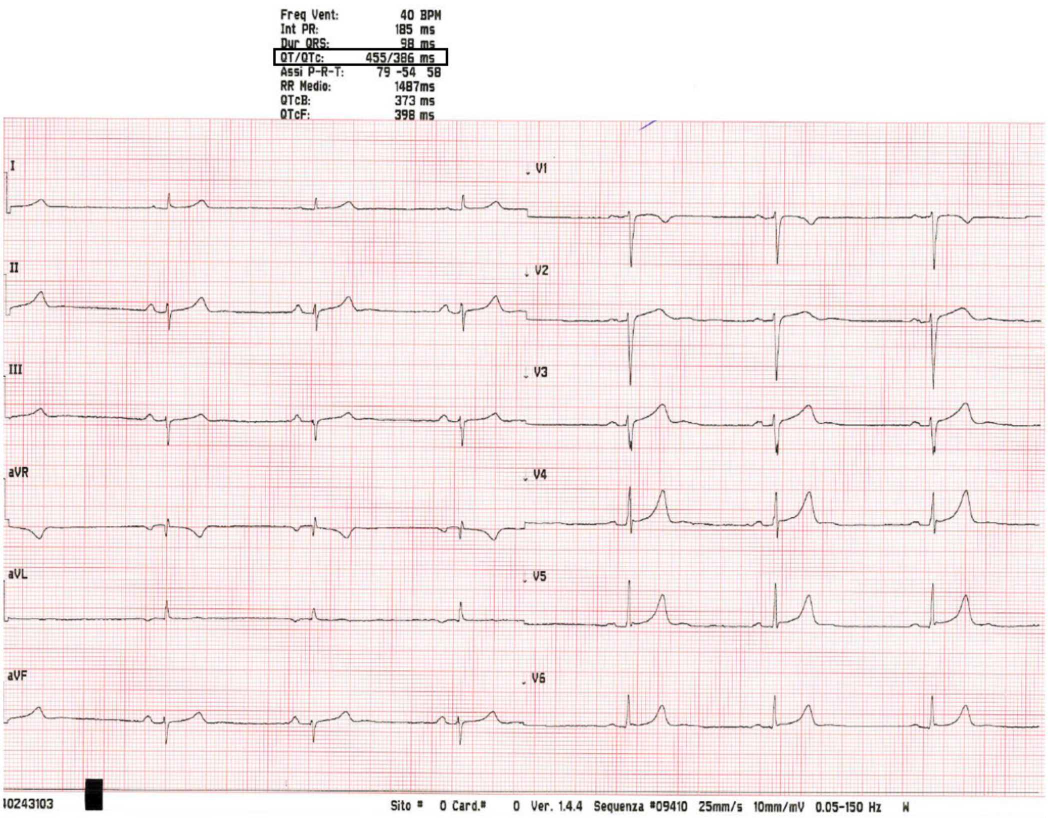

**Supplemental Figure 3a.** Patient n° 7. LQTS female patient (QTc 523 msec, ECG top panel) surviving a previous cardiac arrest and experiencing several appropriate ICD shocks (ICD recording, top panel). Epicardial mapping identified abnormal electrograms (CARTO map, bottom panel), which presented a low amplitude (<1.5mV) and multiple fragmented and double components (bottom panel on the left side) opposed to the normal epicardial signal which is sharp and not fractionated (bottom panel on the right). The distribution of abnormal signals clustered from the epicardial outflow-tract to the infero-lateral peritricuspid region including anterior wall. These abnormalities accounted for a surface area of 19 $cm^2$. The map is rotated towards the left ventricle to show the LV EGM characteristics.

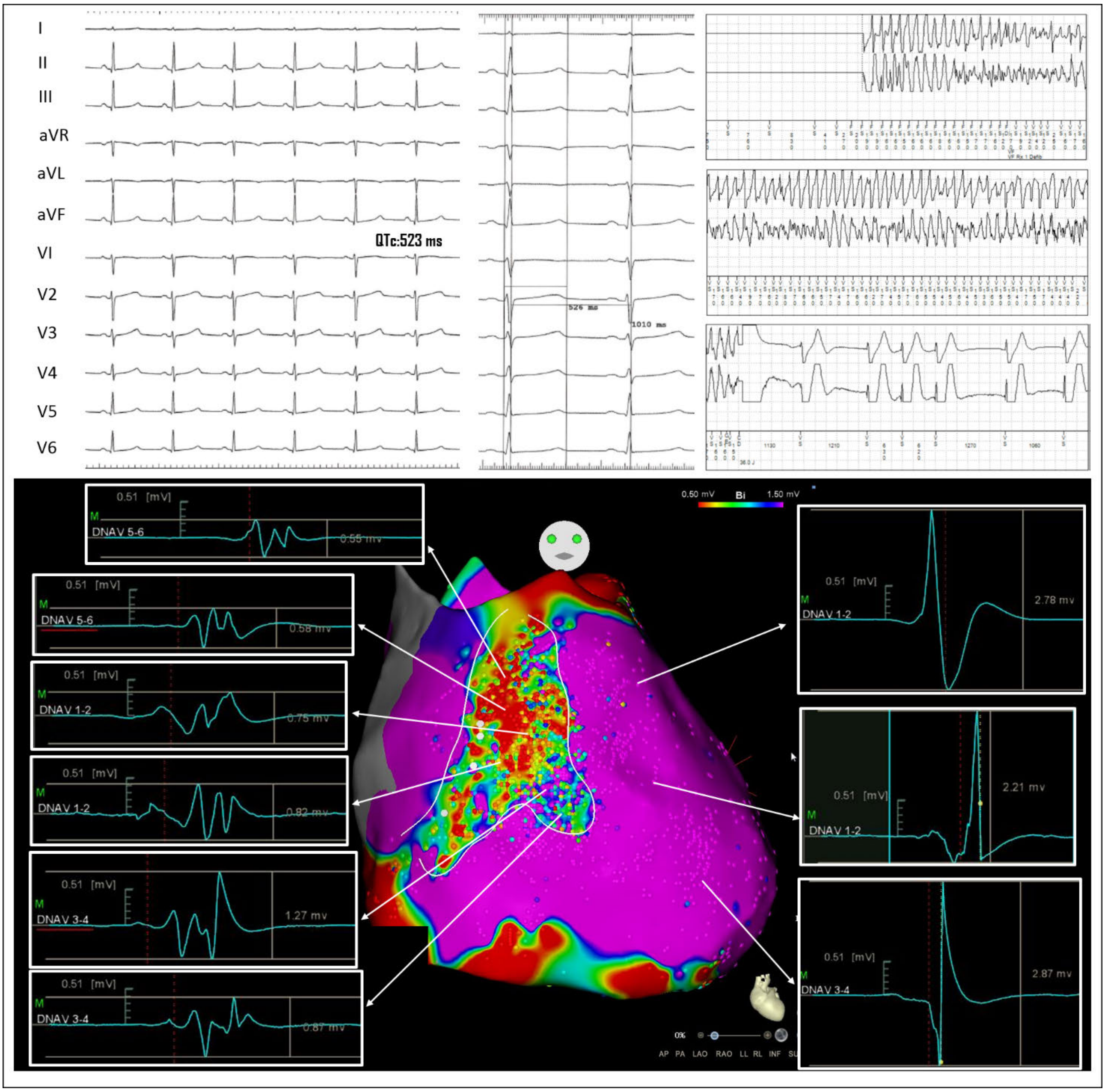

**Supplemental Figure 3b.** Same patient as in Figure 2a. The 12-lead ECG, recorded at the end (left panel) of the procedure, shows QTc shortening (The ECGs are shown at 25 and 50 mm/sec paper speed). Epicardial mapping following catheter ablation demonstrates the disappearance of electrograms fragmentation over the arrhythmogenic substrate area (bottom panel).

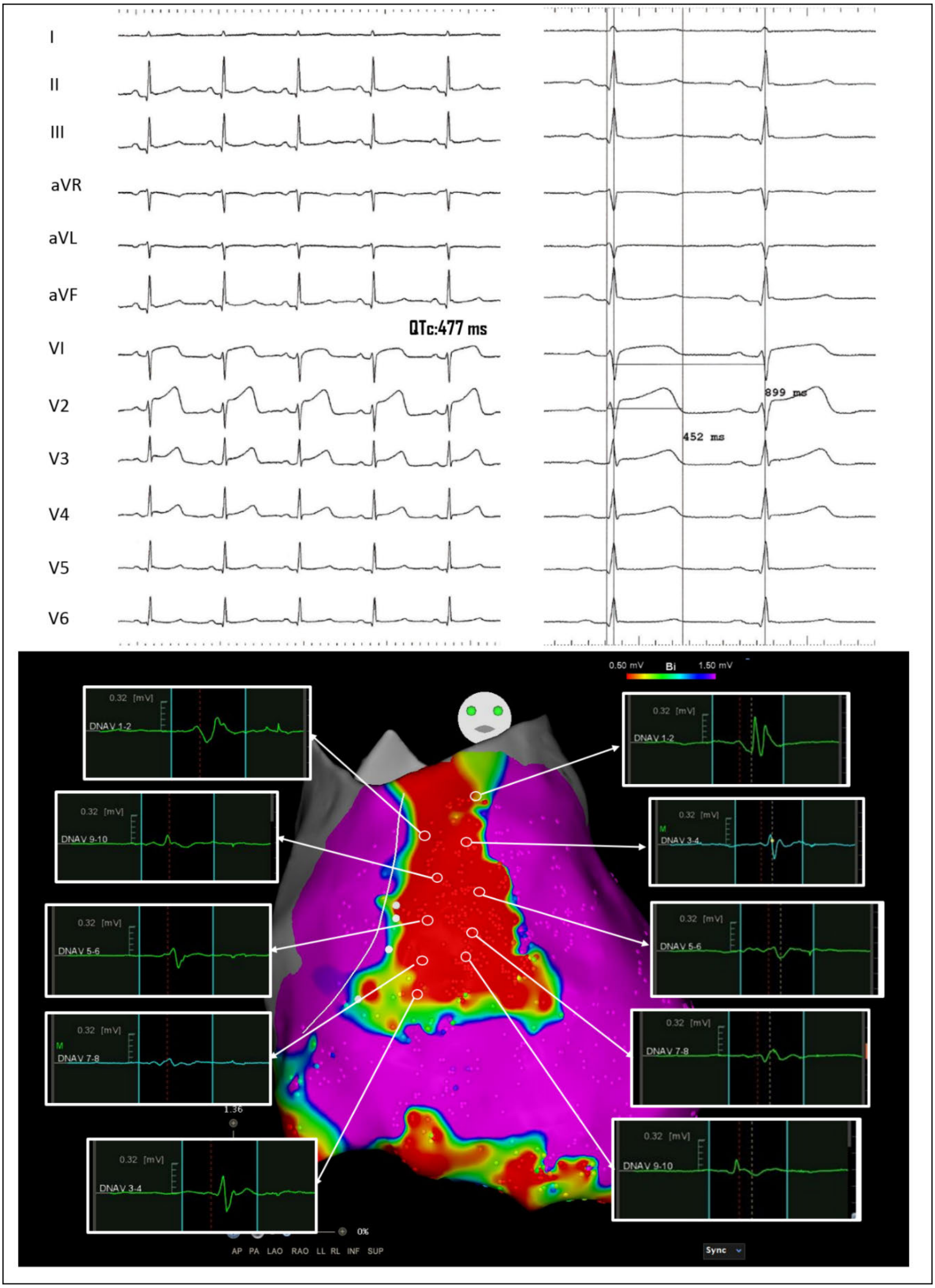

**Supplemental Figure 4a.** Patient n° 8. Patient with LQTS showing prolongation of the QT interval after adrenaline administration (QTc 445 msec at baseline [top panel], 524 msec after adrenaline [bottom panel]). Epicardial mapping identified abnormal electrograms (CARTO map) over the anterior wall of the right ventricle (top panel). The epicardial surface area showing electrical abnormalities increased to 16.6 $cm^2$ (bottom panel on the right).

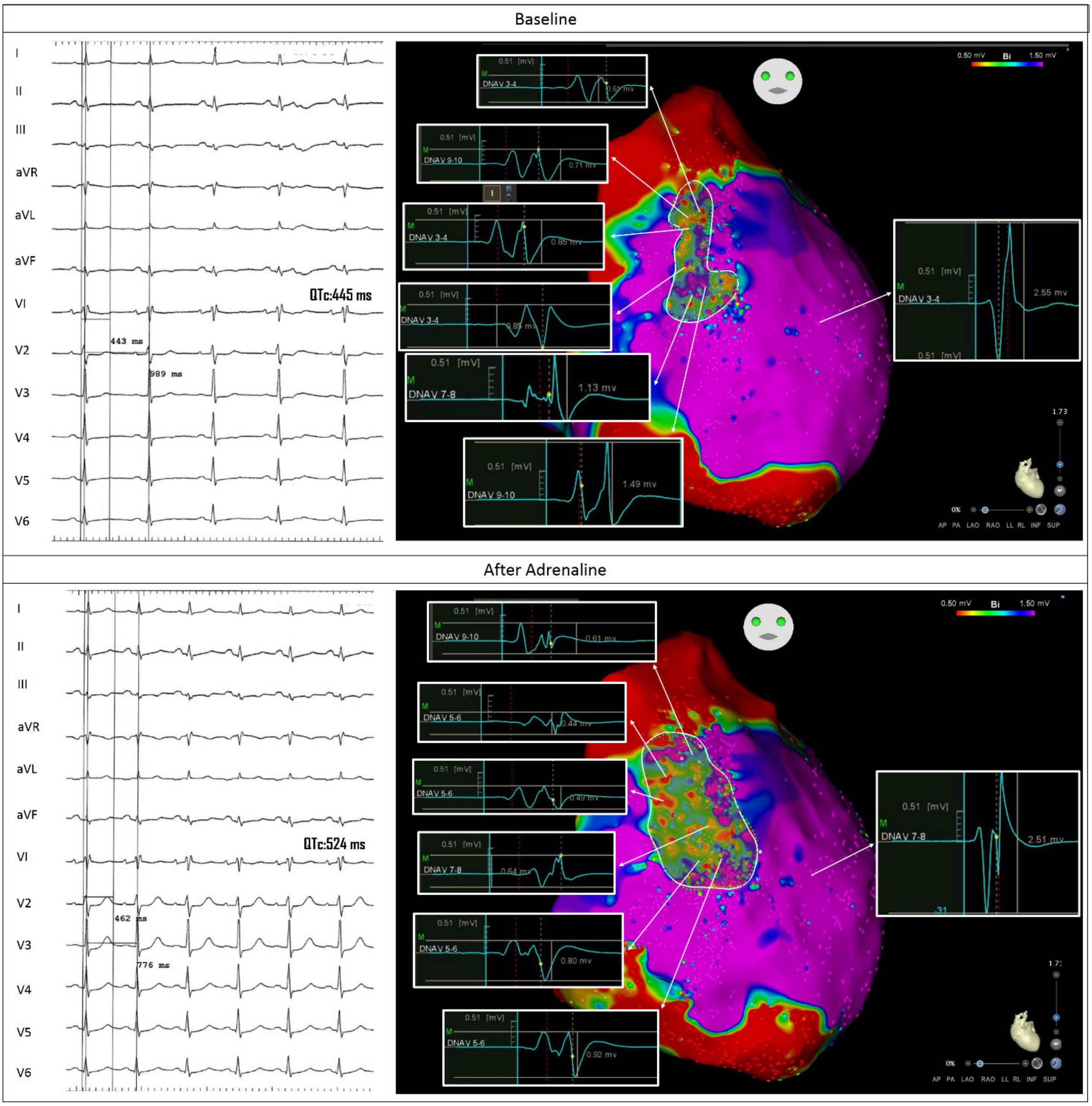

**Supplemental Figure 4b.** The 12-lead ECG shows QTc shortening compared to the baseline conditions (left panel). Epicardial mapping after catheter ablation over the arrhythmogenic substrate shows the abolition of the delayed and fragmented components (right panel).

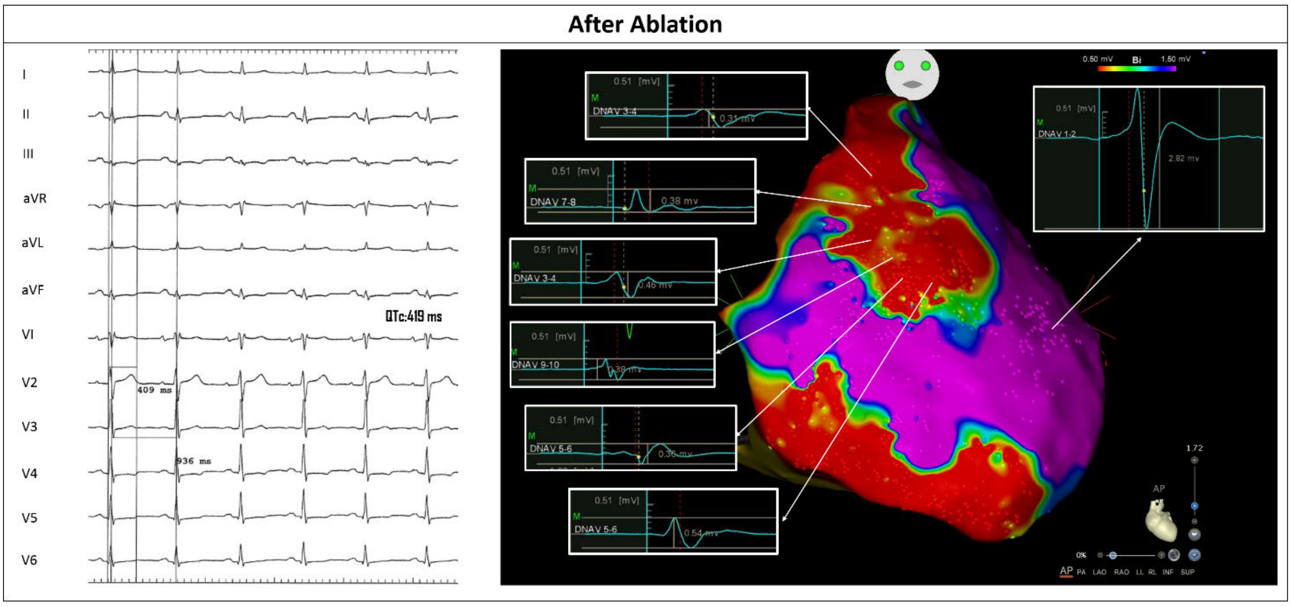

**Supplemental Figure 5a**. Patient n° 8. Patient with LQTS showing prolongation of the QT interval after adrenaline administration (QTc 457 msec at baseline [top panel], 584 msec after adrenaline [bottom panel]). Epicardial mapping identified abnormal electrograms (CARTO map) over the anterior wall of the right ventricle (top panel). The epicardial surface area showing electrical abnormalities (abnormal electrograms are shown in the white boxes) increased to 17.3 $cm^2$ (bottom panel on the right).

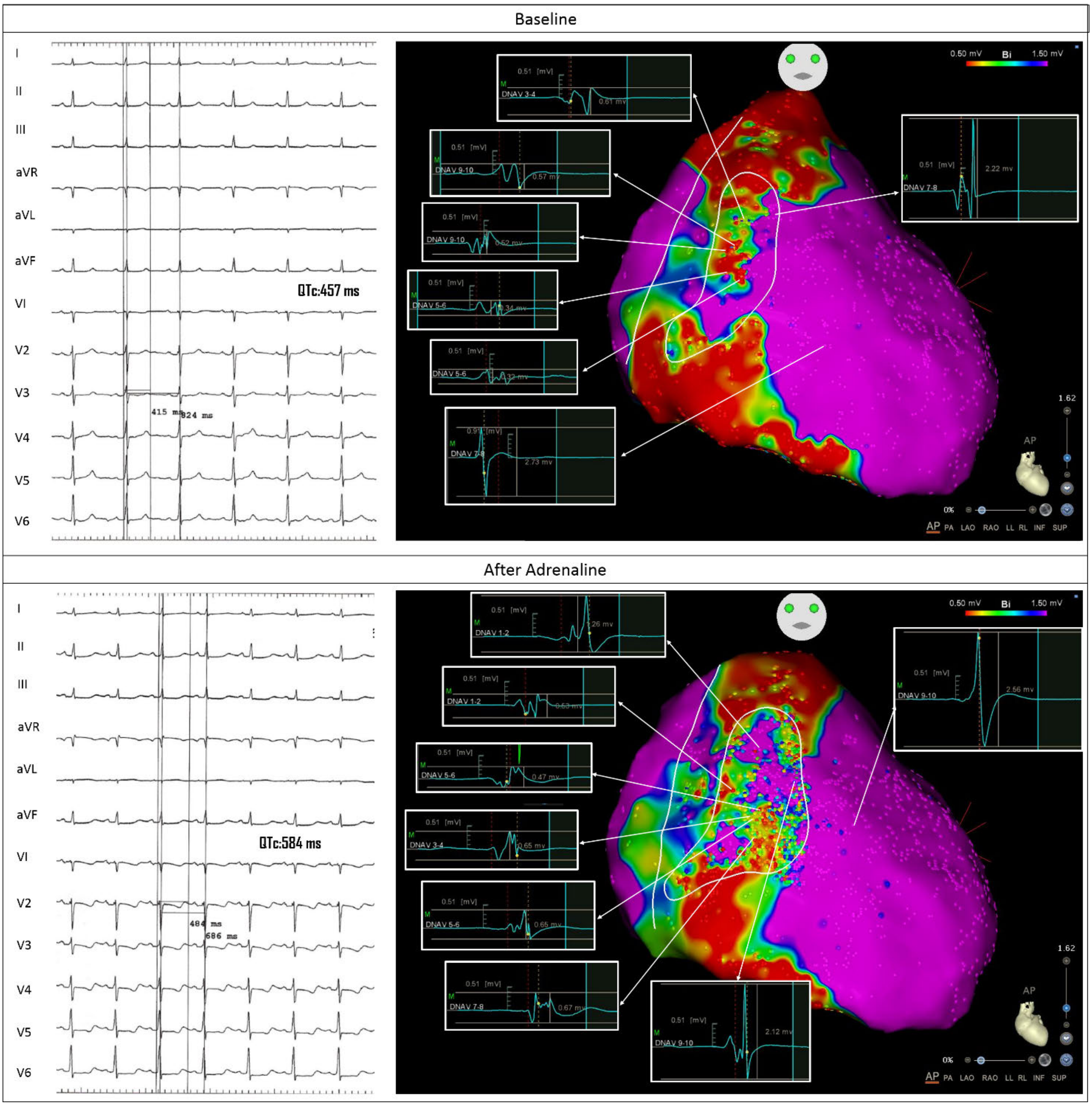

**Supplemental Figure 5b.** The twelve-lead ECG shows mild horizontal ST-segment elevation after ablation. The QTc duration was similar to baseline immediately after the ablation procedure (before 457 vs after ablation 461 msec). Epicardial mapping after catheter ablation over the arrhythmogenic substrate shows the abolition of the delayed and fragmented electrograms (right panel).

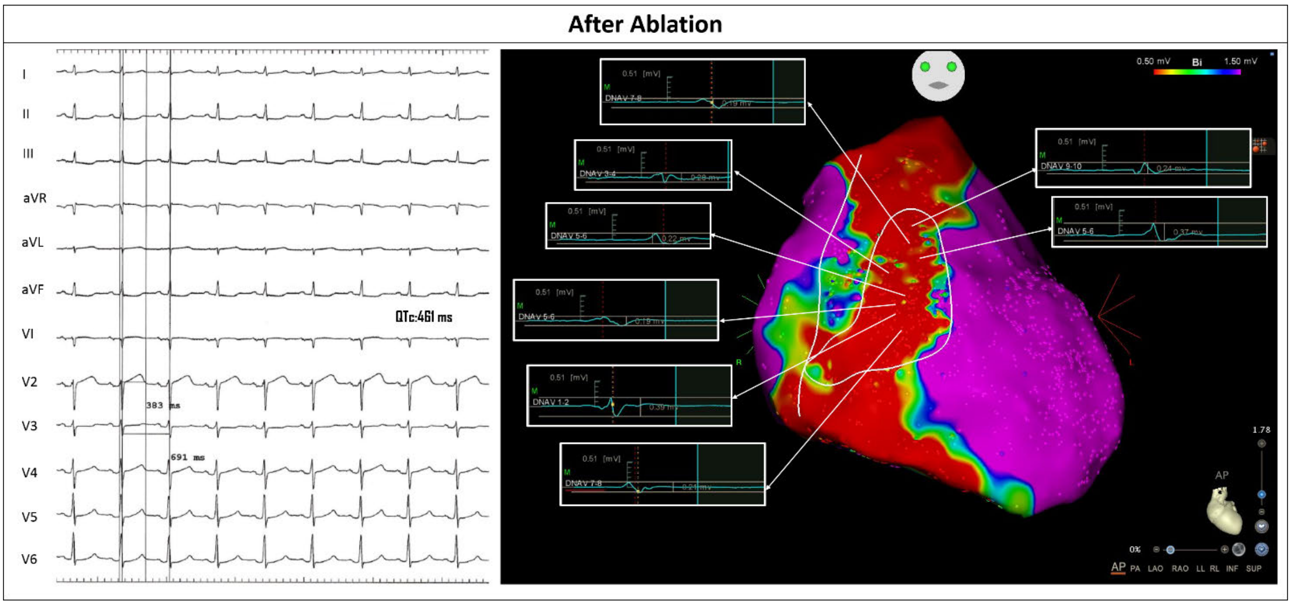

**Supplemental Figure 5c.** Twelve-leads ECG at 3-month follow-up of the same patient as in supplemental figure 5. The QTc interval was shorter compared to the one recorded before the procedure. Leads V1-V3 show resolution of the ST-segment elevation.

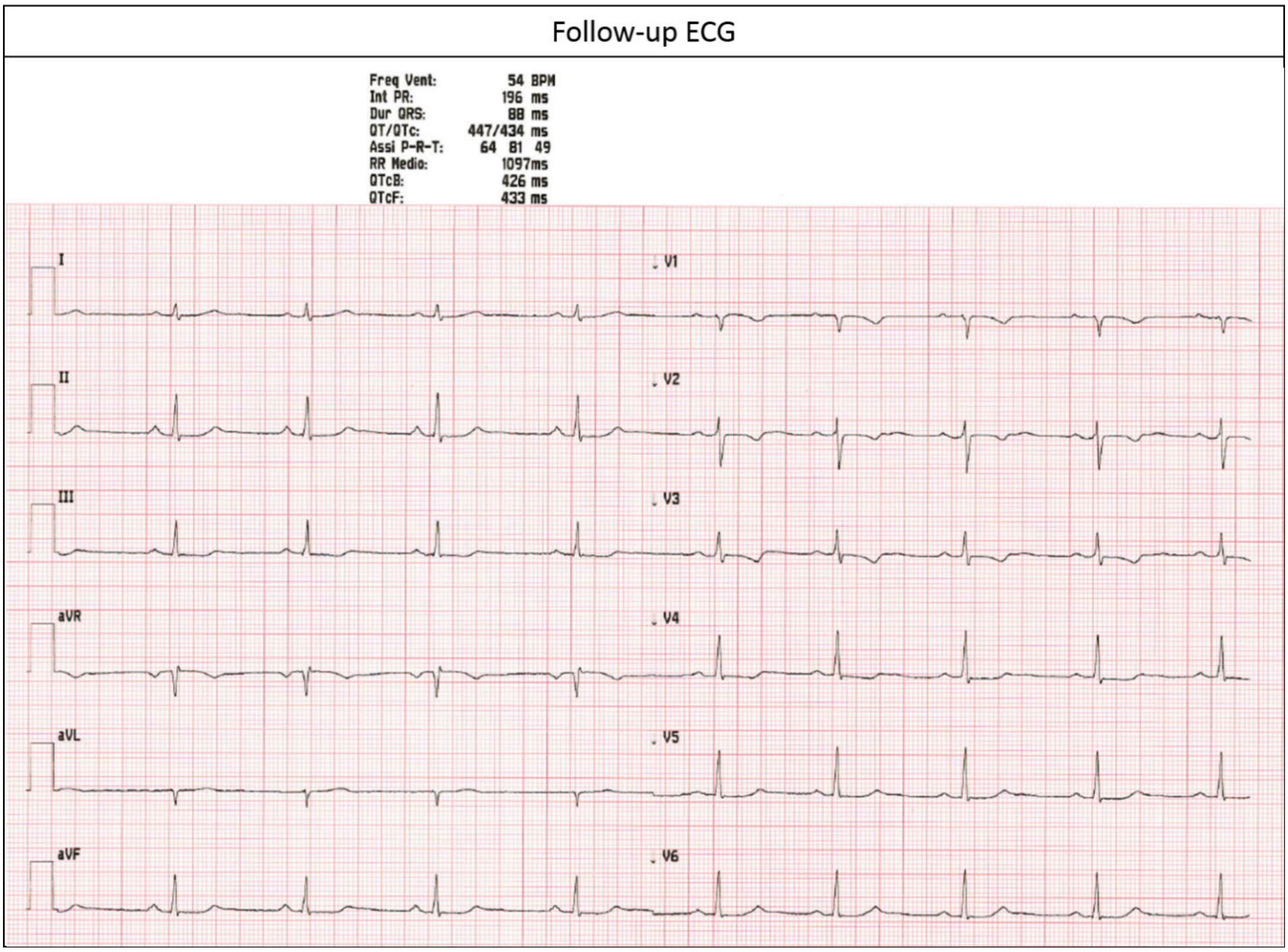

**Supplemental Figure 6**.

Patient n° 5 in table 1 with LQTS (ECG shown in top and middle panels on the left) and Brugada Syndrome (ajmaline test results in middle panel on the left). Epicardial mapping demonstrated an area of fragmented and delayed potentials in the infero-lateral epicardial RV region, whereas typical BrS electrogram fragmentation is shown in the RVOT (top panel). Middle panel shows the results of epicardial mapping after ajmaline infusion with occurrence of the type 1 pattern. The area of abnormalities became confluent. The RVOT region showed typical BrS delayed and prolonged duration electrograms (Brugada area 11.6 $cm^2$); whilst the infero-lateral peritricuspid region maintained the fractionation initially observed. Catheter ablation over these areas resulted in the abolition of electrogram fragmentation, elimination of type 1 pattern and QTc shortening (bottom panel).

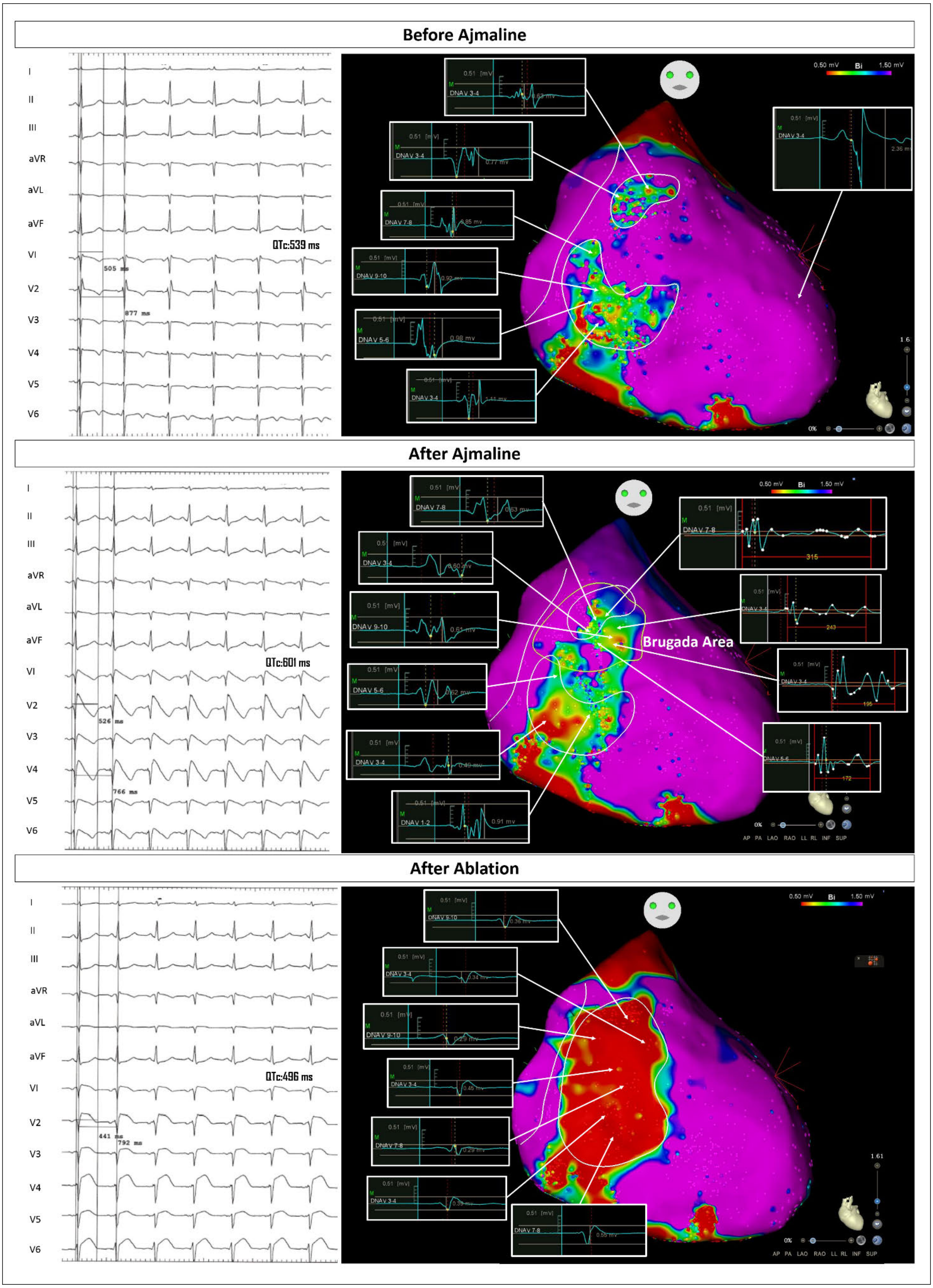

Before Ajmaline
I
II
III
aVR
aVL
aVF
VI
V2
V3
V4
V5
V6
QTc:539 ms
505 ms
877 ms
0.50 mV Bi 1.50 mV
After Ajmaline
QTc:601 ms
526 ms
766 ms
Brugada Area
0.50 mV Bi 1.50 mV
After Ablation
QTc:496 ms
441 ms
792 ms
0.50 mV Bi 1.50 mV

**Supplemental Figure 7**. Patient n° 3 in table 1 with LQTS and ERP in the inferior leads (arrows indicating ER). Top panel shows ECG and epicardial mapping which allowed the identification of an area of fragmented and abnormal potentials in the RVOT and anterior wall. Bottom panel shows the effect of catheter ablation over this area achieving abolition of the fragmentation and QTc shortening.

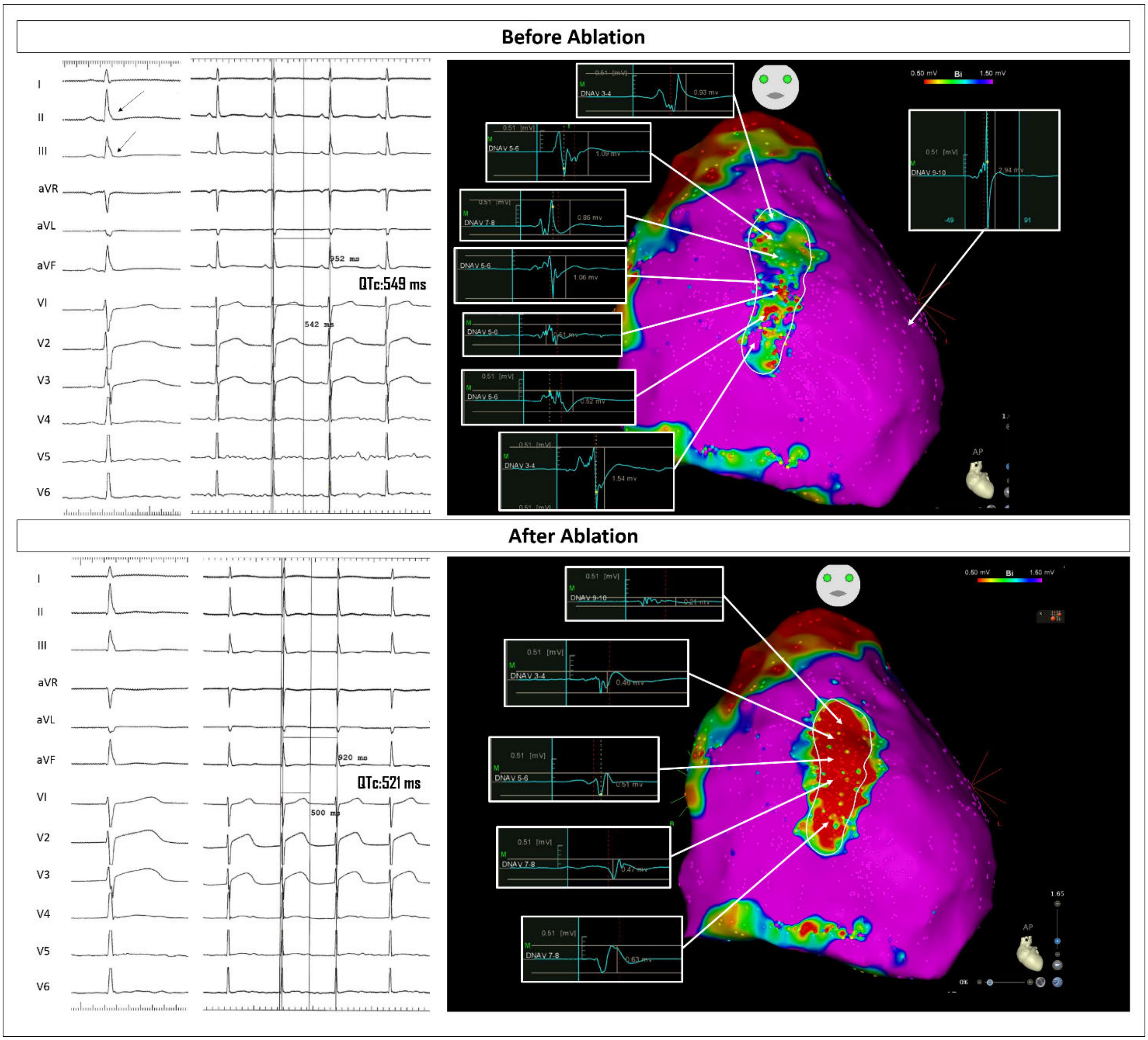

**Supplemental Figure 8**. Twelve-leads ECG at 36 months follow up of same patient as in figures 2 and 3 of the main manuscript (Patient n° 6 in table 1). The QTc interval was shorter compared to the index procedure as well as the ST-segment elevation in V1-V3 completely resolved.

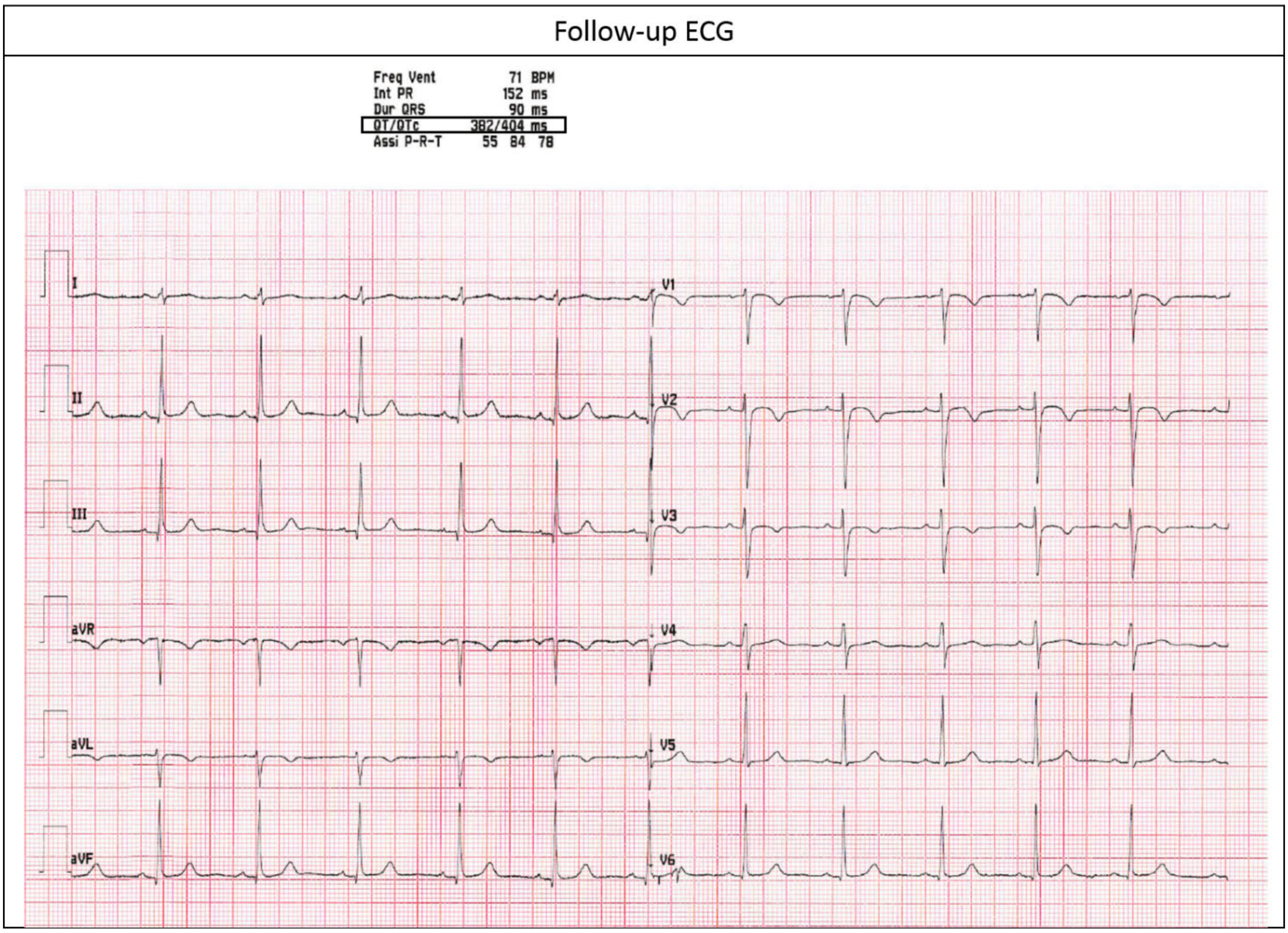

**Supplemental Figure 9**. Example of abnormal epicardial ventricular electrograms typically discovered in patients with right ventricular inherited cardiomyopathies (e.g. Brugada, Long-QT and Early Repolarization Syndromes), treated in our institution.

Right panel shows example of epicardial mapping in a LQTS patient (patient # of the present study cohort). The abnormal signals present a low amplitude (<1.5mV) and are characterized by multiple fragmented and double components. The multiple components could be expression of the local conduction delay and line of block expressed by the separation between the different components. The distribution of abnormal signals extended from the epicardial RVOT to the infero-lateral peritricuspid region including the anterior wall. This geometrical distribution could correspond to the autonomic nervous system on the epicardium of the RV, and these electrophysiological features might represent the structural and functional remodeling due to an imbalanced adrenergic stimulation.

Middle panel shows an example of an historical case of BrS patient having undergone epicardial mapping and ablation. The Potential duration map identifies an area exhibiting delayed and prolonged signals. These electrical abnormalities show a concentric 'onion-like' distribution in the RVOT and anterior epicardial wall. Traditionally, the core region displays the most delayed electrical activity (most prolonged signal in DNAV 5-6), with a less prolongation degree at the periphery (less prolonged activity in DNAV 3-4, 1-2 and 7-8; normal signal in DNAV 9-10).

Left panel shows example of an historical ERS patient having undergone epicardial mapping and ablation at our institution, because of recurrent VF episodes. The anterior wall of the epicardial RV exhibits low-amplitude (<1 mV) and fragmented abnormal electrograms. These regions may harbor microstructural abnormalities and fibrosis ultimately causing the low-voltage and the electrograms fractionation.

### ABNORMAL EPICARDIAL VENTRICULAR ELECTROGRAMS

**Long QT Syndrome**

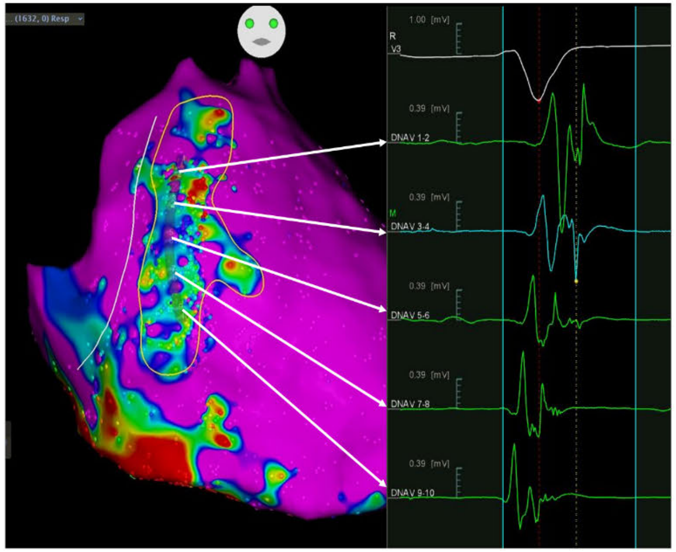


**Brugada Syndrome**

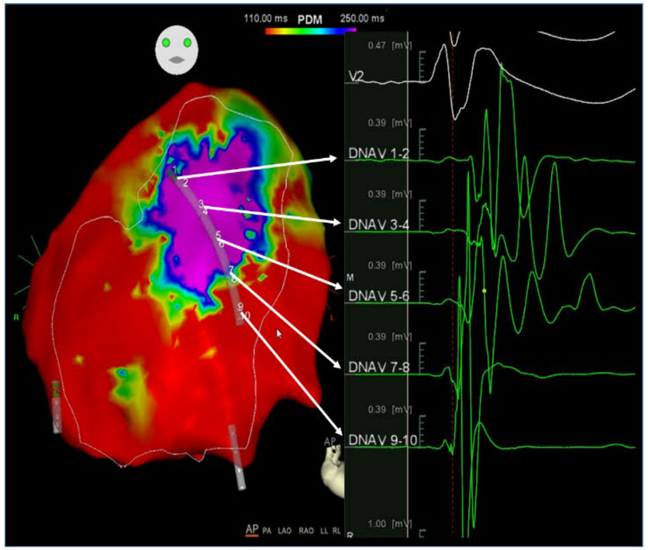


**Early Repolarization Syndrome**

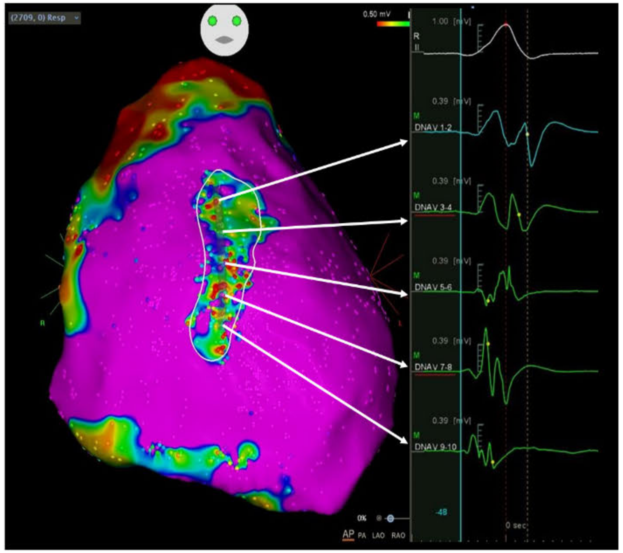
